# Incorporating continuous mammographic density into the BOADICEA breast cancer risk prediction model

**DOI:** 10.1101/2025.02.14.25322305

**Authors:** Lorenzo Ficorella, Mikael Eriksson, Kamila Czene, Goska Leslie, Xin Yang, Tim Carver, Adam E. Stokes, Douglas F. Easton, Per Hall, Antonis C. Antoniou

## Abstract

**Background:** BOADICEA (v7) predicts future breast cancer (BC) risk using data on cancer family history, genetic markers, questionnaire-based risk factors and mammographic density (MD) measured using the 4-category BI-RADS classification. However, BI-RADS requires manual reading, which is impractical on a large scale and may cause information loss.

**Methods:** We extended BOADICEA to incorporate continuous MD measurements, calculated using the automated Volpara and STRATUS software. We used data from the KARMA cohort (60,276 participants; 1,167 incident BC). Associations between MD measurements and BC risk were estimated in a randomly selected training subset (two-thirds of the dataset). Percent MD residuals were calculated after regressing on age at mammography and BMI. Hazard ratios (HRs) were estimated using a Cox proportional hazards model, adjusting for family history and BOADICEA risk factors, and were incorporated into BOADICEA. The remaining one-third of the cohort was used to assess the performance of the extended BOADICEA (v 7.2) in predicting 5-year risks.

**Results:** The BC HRs per SD of residual STRATUS density were estimated to be 1.48 (95%CI: 1.33-1.64) and 1.41 (95%CI: 1.27-1.56) for pre- and post-menopausal women, respectively. The corresponding estimates for Volpara density were 1.27 (95%CI: 1.15-1.40) and 1.38 (95%CI: 1.25-1.54). The extended BOADICEA showed improved discrimination in the testing dataset over using BIRADS, with a 1-4% increase in AUC across different combinations of risk factors. Based on 5-year BC risk with MD as the sole input, approximately 11% of the women were reclassified into lower risk categories and 18% into higher risk categories using the extended model.

**Conclusion:** Incorporating continuous MD measurements into BOADICEA enhances breast cancer risk stratification and facilitates the use of automated MD measures for risk prediction.

## Introduction

Breast cancer (BC) is the most common cancer in women^1,2^. Early detection through screening and risk reduction through preventative medication or surgery are available^3–5^, but these may be associated with overdiagnosis, overtreatment or other side-effects. BC risk prediction models can help targeting such approaches to those likely to benefit, thus optimising resource allocation^6^.

BOADICEA (Breast and Ovarian Analysis of Disease Incidence and Carrier Estimation Algorithm) BC risk model v7^7–9^, implemented in the CanRisk tool^10^ can be used to predict BC risk using data on a combination of factors including cancer family history (FH), pathogenic genetic variants, polygenic scores (PGS), and lifestyle, hormonal, and reproductive risk factors (referred to here as questionnaire-based risk factors, QRFs). BOADICEA also includes mammographic density (MD)^8,11^, using the BIRADS density categories^12^.

MD measures the amount of stromal and epithelial tissue in the breast, which appear as brighter regions in mammograms compared to adipose tissue^13^. MD is usually measured in terms of the proportion of dense breast. Higher MD is associated with increased BC risk^14,15^ and MD has been included in several BC risk models (e.g., Lee *et al.*, 2019^11^, Eriksson *et al.*, 2020^16^, Brentnall *et al.*, 2020^17^). MD is influenced by many other factors including age, body mass index (BMI), and menopausal status, and was found to vary by ethnicity^18–20^. The most widely used method for evaluating MD is the four-category BIRADS (Breast Imaging Reporting & Data System)^12^. BIRADS relies on manual reading of images. However, this is time-consuming and is prone to inter- and intra-operator variability^21,22^. Advances in image-based analysis have led to fully automated tools for measuring MD (e.g. STRATUS^23^, Volpara^24^, Quantra^25^). These produce MD estimates on a continuous scale, expressed as a percentage of either image area (PMD) or breast volume (PVD). STRATUS calculates PMD and can be applied to processed images; Volpara calculates PVD and requires raw images.

Using fully automated tools in clinical practice can be more cost-effective at population scale and improve consistency in MD assessment. Moreover, BC risk varies continuously with MD, and use of categorical measures such as BIRADS results in information loss. Continuous MD measures have been shown to be stronger risk predictors than categorical measures, e.g. in terms of log-odds per adjusted standard deviation (OPERA)^26–28^.

Here, we used data from a large prospective cohort of women enrolled in the mammographic screening programme in Sweden to derive risk estimates for continuous MD measured by Volpara and STRATUS software. We extended BOADICEA by incorporating continuous MD into the BC risk model (v7.2) and validated the resulting model in an independent subset of the cohort.

## Materials and Methods

### Study population

The KARolinska Mammography Project for Risk Prediction of Breast Cancer (KARMA) cohort enrolled women from Sweden (age range 40-74) undergoing mammographic screening between 2011 and 2013^29^; participants were followed until December 2019. Study participants provided self-reported first-degree cancer FH and data on QRFs. The first mammogram at study entry was used to measure MD using STRATUS^23^ and Volpara^30^; the mean density scores of the left and right breasts were used. STRATUS scores were used to derive equivalent BIRADS categorical scores computationally (cBIRADS^31^). Data on cancer diagnoses and deaths were obtained from linkages to healthcare registers.

Genome-wide genotyping data were available for a subset of participants included in previous genotyping experiments^32^, which were used to construct the 313-SNP PGS model^33^. PGS were available for about a quarter of the participants: half of the BC incident cases and a randomly selected subset of unaffected women from the entire cohort.

Analysis was restricted to women who were unaffected with BC at recruitment, had not had risk-reducing mastectomy, and for whom MD measurements were available. To exclude prevalent cancers, women diagnosed with cancer within 1 year from study entry were also excluded. The eligible cohort was split randomly into a *training* (two-thirds of the dataset) and a *testing set* (one third of the dataset), preserving the same case-to-control ratio in both sets. Missing data were imputed using MICE (Multiple Imputation by Chained Equations^34^, Supplementary Materials).

### Statistical Methods

#### Residuals and normalisation

A key assumption of the model is that, by the regressing out the effect of age, the standardised PMD/PVD can then be considered as a fixed covariate. Therefore, we first calculated the residuals of PMD (STRATUS) and PVD (Volpara) after regressing on age at entry and BMI (without interaction) using linear regression (*lm* function, R^35^). Residuals were then transformed through a two-parameter Box-Cox transformation to obtain a gaussian distribution and then further standardised to obtain a standard normal distribution (Supplementary Materials and **Table S1**). Separately, residuals of PMD and PVD were calculated after regressing on age at entry only; residuals were then transformed and standardised in the same way (Supplementary Materials and **Table S1**).

#### Associations with breast cancer risk

Associations between the standardised PMD and PVD residuals and BC risk were evaluated in the training dataset using a Cox proportional hazard model to estimate the hazard ratio (HR) for BC per standard deviation of the standardised residual PMD/PVD. Participants were followed from study entry until the first event among: cancer (in situ/invasive BC, other cancer), prophylactic mastectomy, death, 80 years of age, or last follow-up date. Participants were excluded if censored within 1 year after study entry. Women were considered affected only if they developed invasive BC.

The Cox regression models (*coxph* function, R *survival* package^36^) were adjusted for FH and for the additive effects (on log-scale) of all other BOADICEA QRFs^11^. FH was included as categorical variable based on the number of affected first-degree relatives (0,1, ≥2). An interaction term between menopausal status and standardised residual PMD/PVD was added as a covariate, allowing separate HR estimates for premenopausal and postmenopausal women to be derived.

Analyses were repeated on the subcohort of participants with PGS using a weighted Cox-regression approach; each study participant was weighted by the inverse of their probability of inclusion in the subcohort with genotype data (Supplementary Materials). To accommodate the fact that BMI may not always be available, additional analyses were conducted, based on residuals of PMD and PVD after regressing on age at entry only (Supplementary Materials).

### Augmented BOADICEA

The resulting BC HR estimates for residual STRATUS PMD and Volpara VPD were included in BOADICEA v7 by adding the log(HR) to the linear predictor, thus providing two alternative options for inputting MD in addition to BIRADS. For this, we adopted the methodology previously developed for incorporating continuous QRFs into BOADICEA^7^. Separately, we also updated the model to employ age-specific distributions (before and after 50 years) for BIRADS categories. Details are described in the Supplementary Materials. The extended model is referred to as BOADICEA v7.2.

### Model validation

We used the BOADICEA BC risk models (v7 and v7.2) to predict BC risk in the testing dataset, starting from 1 year after the age at baseline^9^ using Swedish age- and calendar period-specific population cancer incidences. BC risk was calculated at 5 years or the censored age (whichever occurred first); women were considered as unaffected if they developed BC after the predicted 5-year interval.

We investigated the reclassification between risk categories when predicting 5-year BC risks using the two models. Four risk categories were considered, as reported in literature for other risk prediction models^37^: below 1%, between 1% and 1.66%, between 1.66% and 3%, above 3%. To investigate model behaviour in low-risk subpopulations, the lowest risk category was further split at 0.35%, (approximately half the 5-year population risk for a 40y old woman), thus yielding five risk categories. Risks were predicted using MD measures only (to highlight the effect of the different MD reporting methods) or the full available input. Net Reclassification Improvement index (absolute NRI^38^, range [-2,2]) was also calculated. We further assessed the model performance (calibration and discriminative ability) under different risk factor combinations (Supplementary Materials).

## Results

After exclusions, the final dataset (**Tables S2, S3**) included 60,276 women (aged 25-74 years) unaffected at study-entry, 1,167 of whom developed invasive BC (mean/median follow-up of 7.6 years). Of these, 981 women developed invasive BC in the 5-year risk prediction horizon used in the validation stage.

The training set (**Table S4**) comprised 40,184 participants and 778 incident cases of invasive BCs (658 within the 5-year period), whereas the testing set (**Table S5**) comprised 20,092 participants and 389 incident cases. There were no significant differences in the risk factor distributions between the training and testing sets (all *p*-values>0.2). Approximately a quarter of participants in each set had PGS data. Supplementary Materials and **Figure S1** show the distribution of density residuals after regressing on age at entry and BMI.

### Associations with breast cancer risk

**Table 1** shows the estimated HRs per standard deviation (SD) of the standardised STRATUS PMD and Volpara VPD residuals after regressing on age and BMI. When adjusting for all QRFs in BOADICEA except PGS, the HRs per SD of residual STRATUS PMD were estimated to be 1.48 (95%CI: 1.33-1.64) and 1.41 (95%CI: 1.27-1.56) for pre- and post-menopausal women, respectively. The corresponding HR estimates per SD of residual Volpara VPD density were 1.27 (95%CI: 1.15-1.40) and 1.38 (95%CI: 1.25-1.54) for pre- and post-menopausal women, respectively. In analyses based on residuals after regression on age only, the HRs were all slightly (7-9%) larger (**Table S6**).

**Table 1:** Hazard ratio estimates per standard deviation of the standardised STRATUS PMD and Volpara VPD residuals; obtained by regressing on age and BMI, and then by adjusting for FH and QRFs (BMI excluded).

|  | STRATUS |  |  | Volpara |  |  |
| --- | --- | --- | --- | --- | --- | --- |
|  | *Unknown status* | *Pre-menopausal* | *Post-menopausal* | *Unknown status* | *Pre-menopausal* | *Post-menopausal* |
| *Unadjusted for PGS* | 1.44<br>(1.34-1.55) | 1.48<br>(1.33-1.65) | 1.41<br>(1.27-1.56) | 1.32<br>(1.23-1.42) | 1.27<br>(1.15-1.41) | 1.38<br>(1.25-1.54) |
| *Adjusted for PGS* | 1.41<br>(1.31-1.52) | 1.45<br>(1.30-1.62) | 1.38<br>(1.25-1.53) | 1.31<br>(1.22-1.41) | 1.26<br>(1.13-1.39) | 1.37<br>(1.23-1.53) |

In the training subcohort with PGS data, there was a well-fitting linear relationship between HRs estimated with and without adjusting for PGS (*HR_adj_* and *HR_non-adj_*, respectively): for STRATUS, the best fitting linear model was *HR_adj_* = 0.98 · *HR_non-adj_* (Root Mean Squared Error (RMSE) = 0.004); for Volpara, *HR_adj_* = 0.99 · *HR_non-adj_* (RMSE = 0.002). Identical results were obtained when using residuals after regressing on age only. Using those parameters, we calculated the HRs per SD that would have been obtained by adjusting for PGS on the whole training set (**Tables 1** and **S6**). Adjusting for the PGS resulted in an attenuation in the HR estimates by 2% for STRATUS and 1% for Volpara.

### Model implementation

BOADICEA v.7 was extended to incorporate PMD measured using STRATUS, VPD measured using Volpara, and to consider age-specific BIRADS distributions. For this purpose, we used the HRs estimates summarised in **Tables 1** and **S6**, and published BIRADS distributions^8^. All other parameters remained the same as in BOADICEA v7. Since the parameters depend on the availability of information on BMI, PGS and menopausal status, different versions of the model are implemented as follows:

- If STRATUS/Volpara MD are used, the model employs the STRATUS/Volpara HRs estimates from **Table 1** if BMI is provided and STRATUS/Volpara HRs from **Table S6** if BMI is missing; these estimates depend also on menopausal status (pre-, post- or unknown).
- If STRATUS/Volpara MD are used and PGS is available, the PGS-adjusted HR estimates are used.
- If BIRADS is used, BOADICEA selects the parameters specific to the age at risk assessment.

### Risk distributions

#### Predicted range of risks (BOADICEA v7.2)

Figure 1 shows the predicted remaining lifetime BC risks for a hypothetical 40-year-old woman. The range of predicted risks (defined as the 1^st^ to 99^th^ percentile) were similar for STRATUS PMD and Volpara PVD; both were wider than the range of risks obtained using the BIRADS categories. For example, by 80 years, the predicted remaining lifetime BC risks ranged: from 0.041 to 0.148 when using BIRADS; from 0.033 to 0.187 when using STRATUS; and from 0.032 to 0.173 when using Volpara. The risk obtained with BIRADS category A was similar to those at the 20^th^ percentile of STRATUS PMD or Volpara PVD; the risk obtained with BIRADS category D was similar to those at the 95^th^ percentile of STRATUS PMD or Volpara PVD. The predicted remaining lifetime risks by BMI category and MD measurement method are shown in **Supplementary Figures S2-S4**.

**Figure 1:**
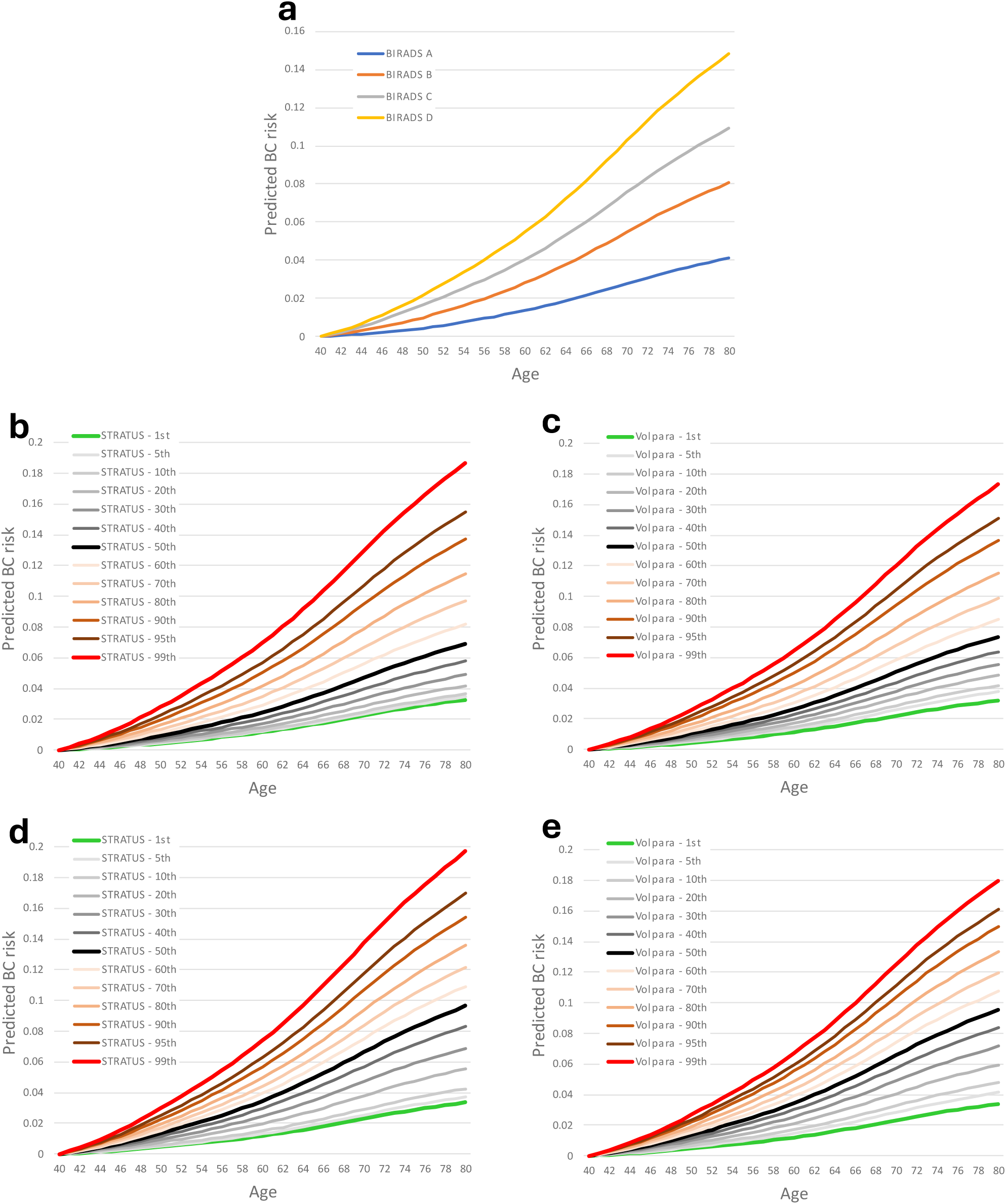
Predicted age-specific BC risk for a 40y woman (born in 2000) to age 80y, using the augmented model. Only MD assumed as input information. Panel **a**, for the 4 different BIRADS categories; panel **b**, for percentiles of the STRATUS PMD distribution in KARMA; panel **c,** for percentiles of the Volpara PVD distribution in KARMA; panel **d**, for percentiles of the STRATUS PMD distribution on women <50 years in KARMA; panel **e,** for percentiles of the Volpara PVD distribution on women <50 years in KARMA. Percentiles considered: 1^st^,5^th^,10^th^ … 90^th^, 95^th^, 99^th^.

#### Predicted 5-year risks in the KARMA testing set

The distribution of predicted 5-year risks in the testing dataset are shown in Figures 2, **S5**, and **S6**. When employing BIRADS categories only, the distributions showed multiple peaks, corresponding to the different BIRADS categories. The risk distributions using STRATUS PMD or Volpara PVD appeared bimodal, reflecting the different HRs per SD for premenopausal and postmenopausal women. However, the distributions became unimodal when additional predictors were included.

**Figure 2:**
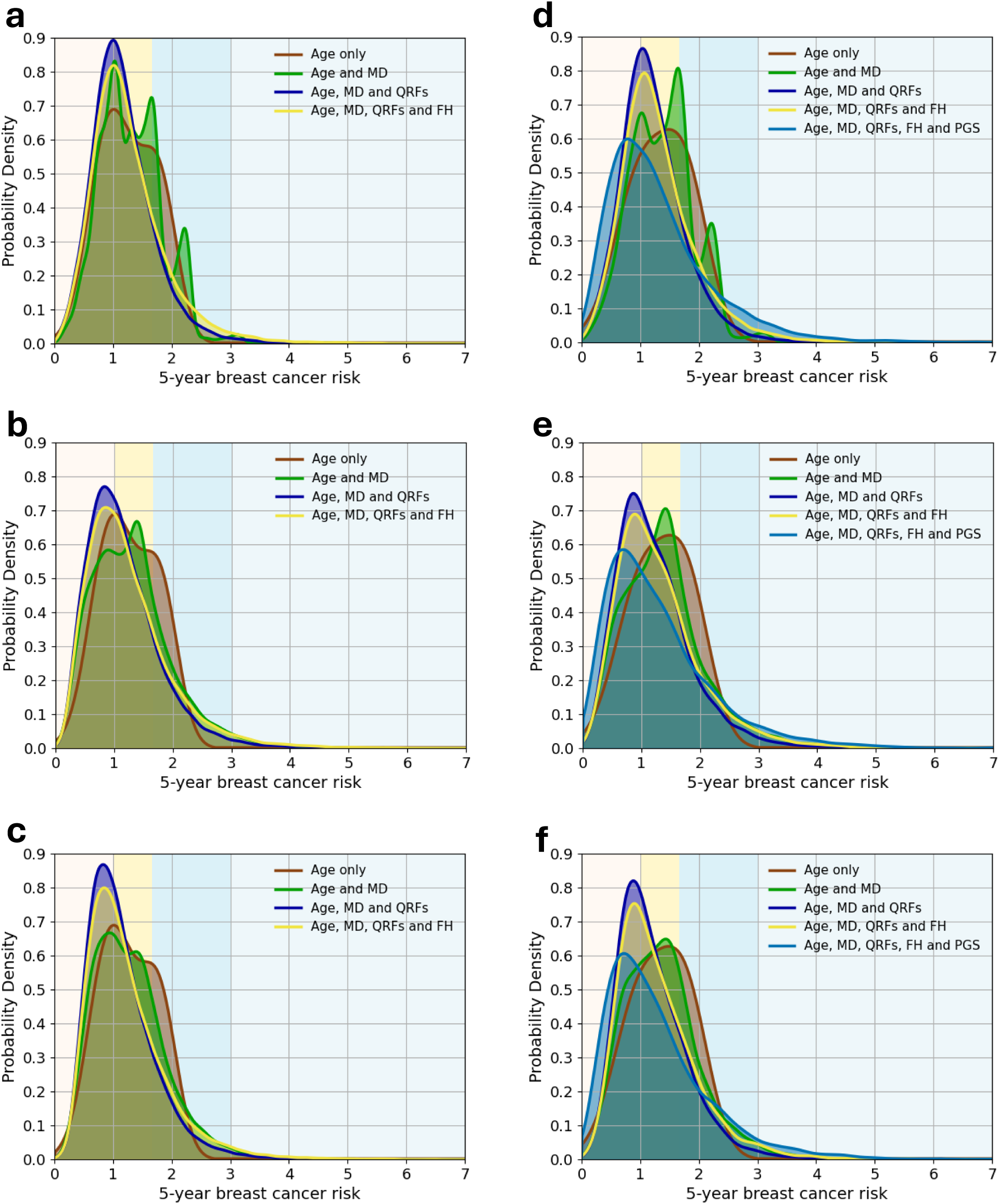
Predicted 5-year BC risk, based on different combinations of risk predictors: (1) age, mammographic density (MD), questionnaire-based risk factors (QRFs), and family history (FH) for all women in the testing dataset (panels **a**, **b**, **c)**; (2) age, MD, QRFs, FH, and polygenic score (PGS) for the subcohort of women with PGS data (panels **d**, **e**, **f)**. All figures show the probability density against the absolute risk: panels (**a**, **d**) using BIRADS categories; panels (**b, e**) using STRATUS PMD measures; panels (**c, f**) using Volpara PVD measures. The backgrounds of the graphs are shaded to indicate four 5-year BC risk categories: less than 1% (light yellow); between 1% and 1.67% (yellow); between 1.67% and 3% (blue); above 3% (light blue).

The predicted risk distributions were slightly wider when using STRATUS and Volpara compared to the distributions obtained using BIRADS; this resulted in differences in risk classification. **Table 2** shows the corresponding proportions of women in the different risk categories. When using MD only, the proportion of women in both the highest and lowest risk categories increased when using continuous MD than BIRADS categories. When using all risk factors (MD, FH, QRFs and PGS), the proportion of women in the highest risk category (≥3%) increased from 0.03 (original model) to 0.04 (Volpara) and 0.05 (age-specific BIRADS, STRATUS). The proportion in the lowest risk category (<0.35%), which was 0.084 in BOADICEA v7, increased when using STRATUS (0.098).

**Table 2:** Classification of women in four risk categories (thresholds derived from Tice *et al.*, 2015^37^); 5-year BC risks calculated using the original BOADICEA v7 model and the augmented model, employing BIRADS categories, STRATUS PMD and Volpara PVD as MD inputs. Risk distributions obtained with models (1) considering MD only; (2) considering MD, FH, QRFs and PGS; All models include age by default.

|  | Distribution, considering MD only |  |  |  |  |  |
| --- | --- | --- | --- | --- | --- | --- |
| **All women** | <0.35% | 0.35%-1% | <1% | 1%-1.67% | 1.67%-3% | >3% |
| *BOADICEA v7* | 0.009 | 0.296 | 0.304 | 0.549 | 0.146 | 0.000 |
| *new model (BIRADS)* | 0.009 | 0.285 | 0.293 | 0.450 | 0.250 | 0.007 |
| *new model (STRATUS)* | 0.030 | 0.324 | 0.354 | 0.399 | 0.224 | 0.022 |
| *new model (Volpara)* | 0.015 | 0.339 | 0.354 | 0.408 | 0.224 | 0.015 |
| **Unaffected only** | <0.35% | 0.35%-1% | <1% | 1%-1.67% | 1.67%-3% | >3% |
| *BOADICEA v7* | 0.009 | 0.298 | 0.307 | 0.549 | 0.145 | 0.000 |
| *new model (BIRADS)* | 0.009 | 0.287 | 0.295 | 0.450 | 0.249 | 0.006 |
| *new model (STRATUS)* | 0.030 | 0.327 | 0.357 | 0.399 | 0.222 | 0.022 |
| *new model (Volpara)* | 0.015 | 0.341 | 0.356 | 0.408 | 0.222 | 0.014 |
| **Affected only** | <0.35% | 0.35%-1% | <1% | 1%-1.67% | 1.67%-3% | >3% |
| *BOADICEA v7* | 0.004 | 0.144 | 0.148 | 0.583 | 0.269 | 0.000 |
| *new model (BIRADS)* | 0.004 | 0.140 | 0.144 | 0.443 | 0.395 | 0.018 |
| *new model (STRATUS)* | 0.018 | 0.146 | 0.164 | 0.413 | 0.392 | 0.031 |
| *new model (Volpara)* | 0.004 | 0.199 | 0.203 | 0.375 | 0.388 | 0.034 |
|  | Distributions, considering full model |  |  |  |  |  |
| **All women** | <0.35% | 0.35%-1% | <1% | 1%-1.67% | 1.67%-3% | >3% |
| *BOADICEA v7* | 0.084 | 0.425 | 0.508 | 0.294 | 0.165 | 0.033 |
| *new model (BIRADS)* | 0.076 | 0.384 | 0.459 | 0.304 | 0.191 | 0.046 |
| *new model (STRATUS)* | 0.098 | 0.397 | 0.495 | 0.272 | 0.183 | 0.050 |
| *new model (Volpara)* | 0.083 | 0.414 | 0.497 | 0.277 | 0.183 | 0.043 |
| **Unaffected only** | <0.35% | 0.35%-1% | <1% | 1%-1.67% | 1.67%-3% | >3% |
| *BOADICEA v7* | 0.084 | 0.427 | 0.512 | 0.293 | 0.163 | 0.033 |
| *new model (BIRADS)* | 0.076 | 0.386 | 0.463 | 0.303 | 0.189 | 0.046 |
| *new model (STRATUS)* | 0.099 | 0.400 | 0.499 | 0.271 | 0.181 | 0.049 |
| *new model (Volpara)* | 0.084 | 0.417 | 0.501 | 0.276 | 0.180 | 0.043 |
| **Affected only** | <0.35% | 0.35%-1% | <1% | 1%-1.67% | 1.67%-3% | >3% |
| *BOADICEA v7* | 0.020 | 0.227 | 0.247 | 0.372 | 0.301 | 0.080 |
| *new model (BIRADS)* | 0.017 | 0.188 | 0.205 | 0.351 | 0.345 | 0.098 |
| *new model (STRATUS)* | 0.013 | 0.199 | 0.212 | 0.309 | 0.347 | 0.132 |
| *new model (Volpara)* | 0.013 | 0.208 | 0.222 | 0.315 | 0.357 | 0.106 |

When using all risk factors (MD, FH, QRFs and PGS) and comparing to the results obtained using BOADICEA v7, the model with age-specific BIRADS resulted in 10% of women reclassified to higher risk categories and none to lower risk categories, with an absolute NRI of 0.021. On the other hand, 8-10% of women were reclassified to lower risk categories and 14% of women towards higher risk categories when STRATUS or Volpara were used (**Table S7**), with absolute NRIs of 0.123 and 0.087 respectively.

### Model validation

The extended model was well calibrated for predicting the 5-year BC risk for all combinations of predictors, showing a slight improvement in the calibration slope compared to BOADICEA v7 (**Table 3**). For example, when using the full input, the calibration slope was 0.98 (95%CI: 0.98-0.99) for STRATUS, Volpara and BIRADS with age-specific categories compared to 0.97 (95%CI: 0.96-0.98) for BOADICEA v7. The corresponding models were also well calibrated in quintiles of predicted risk (Figure 3).

**Figure 3:**
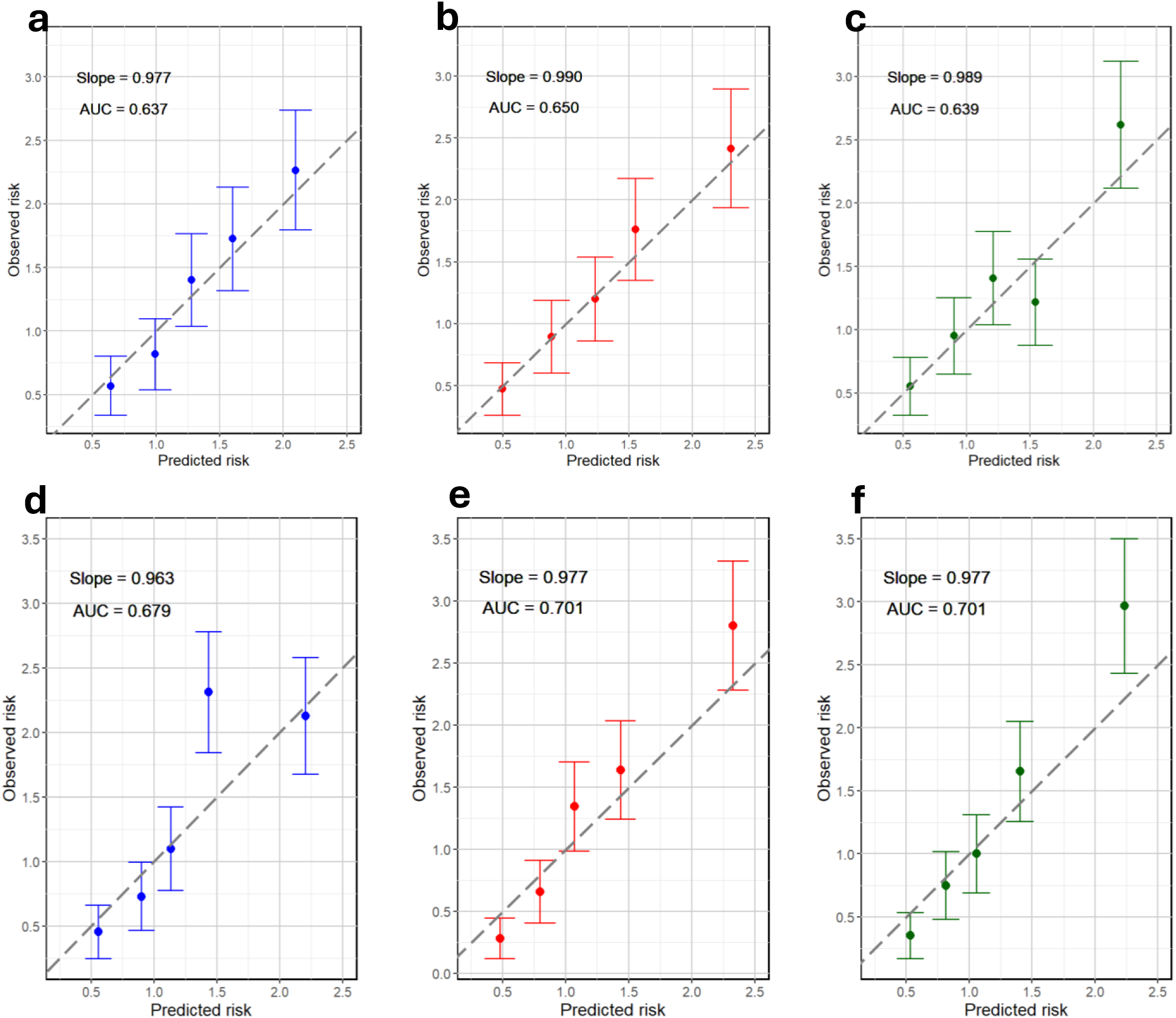
Calibration of the absolute predicted 5-year BC risk, showing the observed and expected risks by quintiles of the predicted risks. Each dot represents the mean observed and predicted risk in the quintile; the bars show the 95% CIs for the observed risks (calculated assuming a Poisson distribution). Panels **a-c**: risks calculated on the entire testing dataset, employing age and MD as inputs. Panels **d-f**: risks calculated on the testing subset containing PGS data, employing all available inputs (age, MD, QRFs, FH and PGS). **a**, **d**: BIRADS; **b**, **e**: STRATUS; **c**, **f**: Volpara.

**Table 3:** Calibration slope and Area Under the Curve (AUC) calculated on risks predicted by the original BOADICEA v7 model and the new model, using BIRADS, STRATUS and Volpara as MD inputs. Multiple inputs combinations tested; 95% confidence intervals reported between parentheses. MD = mammographic density; QRFs = risk factors (all); FH = family history; PGS = polygenic score.

| Inputs combination | Calibration slope on the testing set, full cohort |  |  |  |
| --- | --- | --- | --- | --- |
|  | *BOADICEA v7* | *BIRADS* | *STRATUS* | *Volpara* |
| *age+MD* | 0.98 (0.97 - 0.99) | 1.00 (0.99 - 1.00) | 0.99 (0.98 - 1.00) | 0.99 (0.98 - 1.00) |
| *age+MD+QRFs* | 0.95 (0.94 - 0.96) | 0.97 (0.96 - 0.98) | 0.96 (0.96 - 0.97) | 0.96 (0.95 - 0.97) |
| *age+MD+FH* | 0.99 (0.98 - 1.00) | 1.01 (1.00 - 1.02) | 1.01 (1.00 - 1.02) | 1.01 (1.00 - 1.01) |
| *age+MD+QRFs+FH* | 0.97 (0.96 - 0.98) | 0.98 (0.98 - 0.99) | 0.98 (0.97 - 0.99) | 0.98 (0.97 - 0.99) |
| Inputs combination | Calibration slope on the testing set, subcohort with PGS |  |  |  |
|  | *BOADICEA v7* | *BIRADS* | *STRATUS* | *Volpara* |
| *age+MD* | 0.98 (0.97 - 0.99) | 0.98 (0.97 - 0.99) | 1.00 (0.99 - 1.01) | 0.99 (0.98 - 1.00) |
| *age+MD+QRFs* | 0.95 (0.94 - 0.96) | 0.95 (0.94 - 0.96) | 0.97 (0.96 - 0.98) | 0.96 (0.95 - 0.97) |
| *age+MD+FH* | 1.00 (0.99 - 1.01) | 1.00 (0.99 - 1.00) | 1.02 (1.01 - 1.02) | 1.01 (1.00 - 1.02) |
| *age+MD+QRFs+FH* | 0.97 (0.96 - 0.98) | 0.97 (0.96 - 0.98) | 0.98 (0.98 - 0.99) | 0.98 (0.97 - 0.99) |
| *age+MD+QRFs+FH+PGS* | 0.96 (0.96 - 0.97) | 0.96 (0.95 - 0.97) | 0.98 (0.97 - 0.99) | 0.98 (0.97 - 0.99) |
| Inputs combination | AUC on the testing set, full cohort |  |  |  |
|  | *BOADICEA v7* | *BIRADS* | *STRATUS* | *Volpara* |
| *age+MD* | 0.64 (0.63 - 0.65) | 0.64 (0.63 - 0.65) | 0.65 (0.64 - 0.66) | 0.64 (0.63 - 0.65) |
| *age+MD+QRFs* | 0.65 (0.64 - 0.66) | 0.65 (0.64 - 0.66) | 0.67 (0.66 - 0.68) | 0.66 (0.66 - 0.67) |
| *age+MD+FH* | 0.64 (0.63 - 0.65) | 0.64 (0.63 - 0.66) | 0.66 (0.65 - 0.67) | 0.65 (0.64 - 0.66) |
| *age+MD+QRFs+FH* | 0.66 (0.65 - 0.67) | 0.66 (0.65 - 0.67) | 0.68 (0.67 - 0.69) | 0.67 (0.66 - 0.68) |
| Inputs combination | AUC on the testing set, subcohort with PGS |  |  |  |
|  | *BOADICEA v7* | *BIRADS* | *STRATUS* | *Volpara* |
| *age+MD* | 0.64 (0.63 - 0.66) | 0.65 (0.63 - 0.66) | 0.66 (0.64 - 0.67) | 0.66 (0.64 - 0.67) |
| *age+MD+QRFs* | 0.65 (0.64 - 0.66) | 0.66 (0.65 - 0.67) | 0.68 (0.67 - 0.69) | 0.69 (0.68 - 0.70) |
| *age+MD+FH* | 0.64 (0.63 - 0.66) | 0.65 (0.64 - 0.66) | 0.66 (0.64 - 0.67) | 0.66 (0.64 - 0.67) |
| *age+MD+QRFs+FH* | 0.66 (0.65 - 0.67) | 0.67 (0.65 - 0.68) | 0.69 (0.68 - 0.70) | 0.69 (0.68 - 0.70) |
| *age+MD+QRFs+FH+PGS* | 0.68 (0.67 - 0.69) | 0.68 (0.67 - 0.70) | 0.70 (0.69 - 0.71) | 0.70 (0.69 - 0.71) |

There was an increase in the AUC under the extended models using STRATUS or Volpara, compared to BOADICEA v7, with improvements of 1% to 4% depending on the combinations of predictors used (**Table 3**). For example, when using all predictors including PGS, the AUC was estimated to be 0.70 (95%CI: 0.69-0.71) for STRATUS and Volpara, compared to AUC=0.68 (95%CI: 0.67-0.69) under BOADICEA v7. In contrast, the age-specific BIRADS implementation only improved the AUC slightly, in comparison to BOADICEA v7.

## Discussion

We have extended BOADICEA by incorporating continuous MD. This led to an increased range of predicted BC risks, improved model discrimination and reclassification. The new model was well calibrated in predicting risks across different risk categories.

The estimated relative risks, for women whose PMD/PVD is 1SD higher than the population average (Supplementary Materials and **Table S8**), were consistent with published^16,23,30,39,40^. The 1-2% attenuation of the association between BC and PMD/PVD when adjusting for PGS was also consistent with published estimates^16^. Finally, the OPERA values of our estimates (which here coincide with the estimated HRs per SD) are in line with published OPERA values on continuous MD^26,41,42^.

Incorporating PMD and PVD into BOADICEA allows for additional flexibility in the use of model. Other models incorporate continuous MD data, as either PMD or PVD. For example, the Tyrer-Cuzick model considers Volpara PVD^17^. However, unlike BOADICEA, where MD directly influences the age-specific BC incidence in the model, Tyrer-Cuzick includes Volpara as a post-prediction adjustment to the predicted risk^43–45^. Validation studies of the Tyrer-Cuzick model using Volpara reported AUCs of 0.61-0.62 for the 10-year BC risk (using FH and QRFs, but no genetic data)^44,45^. Other models have been developed to include STRATUS PMD^16,46^ in combination with other AI-based image features; validation studies of those models (in KARMA) reported AUCs of 0.77 for the 2-year BC risk^16^, 0.75-0.65 for the 1 to 10-year BC risks^46,47^.

BMI and MD are not independent variables. However, in practice, MD and BMI might not both be available. To allow for this flexibility, a key aspect of the extended BOADICEA model is that it employs two separate sets of parameters for STRATUS and for Volpara, depending on whether BMI is available. A similar approach is used when PGS data are available, where a somewhat attenuated effect size for MD is assumed based on our estimates (and expected, due to the correlation between MD and PGS).

We also updated the distributions of BIRADS categories for women of age 50 and older. The (small) positive NRI suggests that the previous model slightly (but systematically) underestimated BC risk in women of age 50 and older.

### Strengths

A key strength of the work is the use of the KARMA cohort, which includes information on all the risk factors used in BOADICEA. This has enabled us to estimate the associations of MD with BC risk, while adjusting for all other risk factors included in BOADICEA.

Our approach of first calculating standardised residuals of the continuous MD (Supplementary Materials) ensures that the process can be applied to any combination of age, BMI and PMD/PVD. In addition to the gain in discrimination, a significant advantage of using continuous rather than categorical density is that, by regressing out age, MD can be treated as a fixed covariate, removing the complexities of modelling a time-dependent covariate. The methodology presented here is flexible and can be used, in principle, for any continuous measure that can be suitably standardised. This should allow BOADICEA to be extended to incorporate not only other measures of MD but also other image-based risk measures that are rapidly emerging through novel machine learning approaches^48,49^.

### Limitations

The study has also some limitations. The KARMA dataset had PGS data for about 25% of its participant. Datasets with more complete genetic data could provide more precise estimates on the associations between MD measurements and BC risk after adjusting for PGS. Also, we did not consider continuous MD measures in the context of pathogenic variants (PV) in risk genes: BOADICEA considers PVs, but the data on PVs in KARMA were too sparse to evaluate the model in PV carriers; larger cohorts with PV and MD data are required.

The model validation was performed using a ‘testing’ subset of the cohort that was not employed in the HR estimation; while this subset was independent, it still came from the same population (hence, similar characteristics) of the ‘training’ dataset (**Tables S4, S5**). The performance of the model may therefore have been overestimated, and additional studies are required to assess the model performance in completely different cohorts. On the other hand, the KARMA dataset only reported computationally-derived categorical BIRADS scores; this possibly masked the extent of the model improvement obtained using continuous MD measures (compared to manually-assigned BIRADS scores).

Moreover, since the youngest women in the KARMA cohort were 40 years old at entry, the model was not directly validated for younger ages. Finally, the KARMA cohort is largely composed of White European women; therefore, it less clear whether the estimated parameters would be directly applicable to other ethnic groups. Published data suggest good agreement in the BIRADS distributions for White, Black and Mixed ethnicities; however, the BIRADS distributions for women of Asian ethnicity are markedly different^8,50^. Therefore, it is likely that model parameters for continuous MD should be recalculated, and the model reevaluated, for Asians.

### Concluding remarks

The updated BOADICEA model (v 7.2), incorporating continuous MD measurements alongside the BIRADS classification, is available online through CanRisk (www.canrisk.org^10^). Although the routine availability of continuous MD measures varies by country, the updated model provides a means of facilitating population-scalable approaches to incorporating MD into routine BC risk assessment.

## Data Availability

Data from the KARMA study are available upon request from Karolinska Institutet, through the MTA form available at karmastudy.org/contact/data-access/

## Notes

**Funding information** This work was supported by Cancer Research UK grant: PPRPGM-Nov20\100002. ACA is supported by Cancer Research UK grant: SEBCD3-2024/100001.

### Competing Interest Statement

LF, TC, DFE and ACA are listed as creators of the BOADICEA model, which has been licensed by Cambridge Enterprise (University of Cambridge).

### Funding Statement

This work was supported by Cancer Research UK grant: PPRPGM-Nov20\100002. ACA is supported by Cancer Research UK grant: SEBCD3-2024/100001.

### Author Declarations

KARMA study was approved by the ethics review board at Karolinska Institutet. All participants provided written informed consent to participate in the study before taking part.

