## Supplementary Materials for "Incorporating continuous mammographic density into the BOADICEA breast cancer risk prediction model"

#### Materials and Methods

##### Imputation

Completeness for individual risk factors ranged from 68% (93% if excluding menopausal status/age at menopause) to 94%, and only ~60% of the participants had complete data for all risk factors (~84% if excluding menopausal status/age at menopause). We imputed missing data using MICE (Multiple Imputation by Chained Equations[1]). The imputation equations were developed solely on the training set and separately applied to both the training and the testing set, ensuring independence between the training and validation steps. We generated 10 imputed sets with 20 successive iterations each. Downstream steps were performed on each imputed set separately. Results were then pooled together by applying Rubin's rules[2].

For each imputed set, the training and testing subsets were compared by applying Pearson's chi-squared test to categorical variables and a Kolmogorov-Smirnov test to continuous variables.

##### Statistical Methods

*Residuals calculation*

Residuals of PMD (STRATUS) and PVD (Volpara) after regressing on age at entry and BMI (without interaction) were calculated using linear regression (*lm* function in R[3]).

$$X = MD - \lambda_0 - \lambda_1 \cdot age - \lambda_2(BMI)$$

where $X$ are the residuals after regressing, MD is the mammographic density (PMD or PVD), age (in years) is treated as a continuous variable and BMI is treated as a categorical one (according to its use in BOADICEA). Residuals were also calculated after regressing on age at entry only:

$$X = MD - \lambda_0 + \lambda_1 \cdot age.$$

Regression parameters are shown in **Table S1**.

*Normalisation*

Residuals were transformed through a two-parameter Box-Cox transformation to obtain a gaussian distribution (step 1), and then further standardised to obtain a standard normal distribution (step 2):

$$X' = \frac{(X + \lambda_0 + 1)^{\lambda_3} - 1}{\lambda_3} \; ; \; X'' = \frac{X' - \mu(X')}{\sigma(X')}$$

where $X$ are the residuals after regressing; $X'$ are the transformed standardised residuals after step 1; $X''$ are the standardised residuals after step 2, employed in all downstream analyses; $\mu$ and $\sigma$ are the mean and standard deviation, respectively.

$\lambda_0$ and $\lambda_3$ are parameters specific for the residuals calculated on this dataset. $\lambda_0$is the intercept of the linear regression that originated the residuals $X$; this guarantees that $X + \lambda_0 + 1 > 0 \; \forall \; X$, i.e. for any possible (user-defined) input. $\lambda_3$was calculated through minimisation of the Kolmogorov-Smirnov test statistic (*ks.test* function in R[3]), comparing the distribution of transformed residuals with a Gaussian distribution having the same mean and standard deviation (*optimise* function in R[3]). Normalisation parameters are shown in **Table S1**.

##### *Associations with breast cancer risk*

Analyses were repeated using the subcohort of participants with PGS. In this dataset, HRs were initially estimated by adjusting only for QRFs and FH, and then further adjusting for PGS. As this subcohort was enriched for incident breast cancers[4], we employed a weighted Cox-regression approach, in which each study participant was weighted by the inverse of their probability of inclusion in the subcohort with genotype data[5,6]. We computed the HRs by combining the results from all imputed datasets, for premenopausal women, postmenopausal women and women with unspecified menopausal status. The PGS-adjusted HRs from this subcohort may not be directly compared with the (unadjusted) HRs from the whole cohort. We therefore performed a linear regression analysis in this subcohort, with the PGS-adjusted HRs as the outcome and the unadjusted HRs as a covariate. The regression parameters were then used to predict the HRs that would have been obtained if we had adjusted for PGS in the whole training set (i.e. if we had had the required PGS data for all participants).

BMI may not always be available in practice (in CanRisk, age at entry is the minimal input required for running the BC risk model); therefore, it might not always be possible to regress on BMI. Hence, we performed additional analyses for which we computed residuals of PMD and PVD after regressing on age at entry only, and transformed the residuals as described above. We then repeated the Cox regression analyses in the whole cohort and the weighted analyses in the subset with PGS data, as above. The only difference in these analyses was that BMI was included as an (optional) adjustment variable in the HR estimation process, together with all other (optional) risk factors.

#### Augmented BOADICEA

##### *Integrating estimated HRs into BOADICEA*

Incorporating continuous risk factors into BOADICEA introduces a computational challenge in deriving the baseline hazard since the number of possible risk factor combinations is effectively infinite. We therefore adopted the methodology previously developed for incorporating continuous QRFs into BOADICEA and used to include height as a risk factor in the model[7].

Briefly, this involves discretising the continuous QRF distribution into a relatively small number $(n + 1)$ of bins. The $n$ bins represent the whole population and can be used for constraining the cancer incidences and therefore computing the baseline rates. The probability mass and relative risk (RR) for each bin are calculated from the probability density and RR function for the continuous QRF (the standard approach for discretising QRFs[8]). The $(n + 1)^{th}$ bin only contains the individual under study, and it is assigned the RR calculated at the measured value; this way, the exact value of the RR appropriate for the MD of the individual can be used. Lee *et al.*[7] shows that this method provides very accurate risk estimates even with relatively small *n*. Here we used *n*=5, as also used for height.

*Distributions of BIRADS categories*

We also updated the model part that incorporates categorical MD. BOADICEA v7 employs age-specific RRs (before and after 50 years); however, it assumes that the distribution across the four BIRADS categories does not vary with age. For each ethnicity in the model (East Asian, South Asian, Black, Mixed, White), distributions are based on women younger than 50 years (from the BCSC dataset[9], analysed in Tice *et al.*, 2008[10] and Ficorella *et al.*, 2025[11]). Here, we modified the model so that it employs age- and ethnicity-specific RRs and distributions. The ethnicity-specific distributions of BIRADS for women of age 50 and older were also obtained in Ficorella *et al.*, 2025[11] using the same BCSC dataset[9].

#### Model validation

We assessed the model performance under various scenarios depending on the available risk factor data (BOADICEA allows for missing information[8]). These included: MD only; MD and QRFs; MD, QRFs, and FH; and MD, QRFs, FH, and PGS. The last scenario was assessed in the testing subset with PGS information, assuming participant weights as outlined above. For each combination, we compared the predicted risks obtained using BOADICEA v7 (assuming BIRADS categories), with those predicted using BOADICEA v 7.2 (BIRADS categories, STRATUS PMD and Volpara VPD).

Model calibration was assessed by calculating the ratio of the expected to the observed BC risk and by calibration plots, comparing observed and expected risks by quantile categories of the predicted risks. We also calculated the calibration slope using logistic regression, by regressing the BC status on (0/1) the log-odds of the predicted risk. Under a perfectly calibrated model, the slope is expected to be equal to 1.

We assessed model discrimination by calculating the Area Under the Receiver operating characteristic curve (AUC). For the full testing set, the AUC was estimated using the Mann-Whitney-Wilcoxon test. When using the subcohort of participants with PGS, the AUC was calculated using the *WeightedAUC* function from the *WeightedROC* R package[12].

### Results

**Figure S1**, shows the distribution of density residuals after regressing on age at entry and BMI (before and after transformation/standardisation), in the whole cohort and in the subset of participants with incident invasive BC. The shape and width of the distribution of residuals prior to standardisation differed between STRATUS and Volpara (PMD and PVD, respectively), with the distribution being wider for STRATUS. As expected, affected women had higher MD (Kolmogorov-Smirnov test, *p*-value $< 10^{-5}$ for BIRADS, *p*-value $< 10^{-16}$ for continuous methods), and the mean of the standardised residual PMD/PVD was higher in affected women (0.33 STRATUS, 0.28 Volpara) compared to the whole dataset (mean = 0), consistent with the association between MD and risk.

The estimated HRs per 1 SD of the standardised residuals of PMD/PVD cannot be directly compared with published data, which report RRs per 1 SD of the PMD/PVD. So, we calculated the RRs for women whose PMD/PVD is 1SD higher than the population average (in the KARMA cohort), for different ages and BMIs (**Table S8**).

### Figure and table legends

**Figure S1:** Distribution of mammographic density (PMD for STRATUS, PVD for Volpara) residuals after regressing on age and BMI. Panel **a**: residuals before Box-Cox transformation and standardisation. Panel **b**: transformed residuals.

**Figure S2:** Predicted age-specific BC risk for a 40y woman (born in 2000) to age 80y, using the augmented model. MD (as BIRADS categories) and BMI assumed as input information. Panel **a**, BMI < 18.5; panel **b**, BMI in [18.5, 25); panel **c**, BMI in [25, 30); panel **d**, BMI ≥ 30.

**Figure S3:** Predicted age-specific BC risk for a 40y woman (born in 2000) to age 80y, using the augmented model. MD (as STRATUS PMD) and BMI assumed as input information. Panel **a**, BMI < 18.5; panel **b**, BMI in [18.5, 25); panel **c**, BMI in [25, 30); panel **d**, BMI ≥ 30. Percentiles considered: $1^{st}$,$5^{th}$,$10^{th}$ – $90^{th}$, $95^{th}$, $99^{th}$ of the STRATUS PMD distribution on women <50 years in KARMA.

**Figure S4:** Predicted age-specific BC risk for a 40y woman (born in 2000) to age 80y, using the augmented model. MD (as Volpara PVD) and BMI assumed as input information. Panel **a**, BMI < 18.5; panel **b**, BMI in [18.5, 25); panel **c**, BMI in [25, 30); panel **d**, BMI ≥ 30. Percentiles considered: $1^{st}$,$5^{th}$,$10^{th}$ – $90^{th}$, $95^{th}$, $99^{th}$ of the Volpara PVD distribution on women <50 years in KARMA.

**Figure S5:** Predicted 5-year BC risk based on different combinations of risk predictors: age, mammographic density (MD), questionnaire-based risk factors (QRFs), and family history (FH). (1) women in the testing dataset who developed invasive BC after entering the study (‘affected’, panels **a**, **b**, **c**); (2) unaffected women in the testing dataset (panels **d**, **e**, **f)**. All figures show the probability density against the absolute risk: panels (**a**, **d**) using BIRADS categories; panels (**b**, **e**) using STRATUS PMD measures, panels (**c**, **f**) using Volpara PVD measures. The backgrounds of the graphs are shaded to indicate four 5-year BC risk categories: less than 1% (light yellow); between 1% and 1.67% (yellow); between 1.67% and 3% (blue); above 3% (light blue).

**Figure S6:** Predicted 5-year BC risk based on different combinations of risk predictors: age, mammographic density (MD), questionnaire-based risk factors (QRFs), family history (FH) and polygenic score (PGS). (1) women in the testing subset with PGS data who developed invasive BC after entering the study (‘affected’, panels **a**, **b**, **c**); (2) unaffected women in the testing subset with PGS data (panels **d**, **e**, **f**). All figures show the probability density against the absolute risk: panels (**a**, **d**) using BIRADS categories, panels (**b**, **e**) using STRATUS PMD measures, panels (**c**, **f**) using Volpara PVD measures. The backgrounds of the graphs are shaded to indicate four risk categories: less than 1% (light yellow); between 1% and 1.67% (yellow); between 1.67% and 3% (blue); above 3% (light blue).

**Table S1**: Regression and standardisation parameters used for calculating residuals of PMD (STRATUS) and PVD (Volpara) and then normalising them to achieve a standard normal distribution. Regression required $\lambda_0$, $\lambda_1$, and $\lambda_2$; normalisation required $\lambda_0$, $\lambda_3$, $\mu(X')$, and $\sigma(X')$.

**Table S2:** KARMA dataset employed in this study before imputing missing values. Risk factor categories as defined in the BOADICEA model. IQR = interquartile range; SD = standard deviation; FDR = first-degree relatives; BC = breast cancer; HRT = hormone replacement therapy.

**Table S3:** KARMA dataset employed in this study after imputing missing values. Risk factor categories as defined in the BOADICEA model. IQR = interquartile range; SD = standard deviation; FDR = first-degree relatives; BC = breast cancer; HRT = hormone replacement therapy.

**Table S4:** Training dataset, randomly extracted (*R* seed= 24) from the imputed KARMA dataset (table S3). Risk factor categories as defined in the BOADICEA model. IQR = interquartile range; SD = standard deviation; FDR = first-degree relatives; BC = breast cancer; HRT = hormone replacement therapy.

**Table S5:** Testing dataset, randomly extracted (*R* seed= 24) from the imputed KARMA dataset (table S3). Risk factor categories as defined in the BOADICEA model. IQR = interquartile range; SD = standard deviation; FDR = first-degree relatives; BC = breast cancer; HRT = hormone replacement therapy.

**Table S6:** Hazard ratio estimates per standard deviation of the standardised STRATUS PMD and Volpara VPD residuals; obtained by regressing on age, and then by adjusting for FH and QRFs (BMI included).

**Table S7:** Summary of the reclassification towards lower, identical, or higher categories when comparing risks predicted with the original BOADICEA v7 model vs the augmented model (using BIRADS categories, STRATUS PMD and Volpara PVD as MD inputs. Thresholds of risk categories derived from Tice *et al.*, 2015[13]. Risk distributions obtained with models (1) considering MD only; (2) considering MD, FH, QRFs and PGS. All models include age by default.

**Table S8:** Hazard ratio estimates for a woman with STRATUS PMD or Volpara VPD 1SD higher than the population average (in the KARMA cohort); calculated for different ages and BMI categories. Women of age 40y and 50y were assumed to be pre-menopausal; women of age 60y and 70y were assumed to be post-menopausal.

### Figures

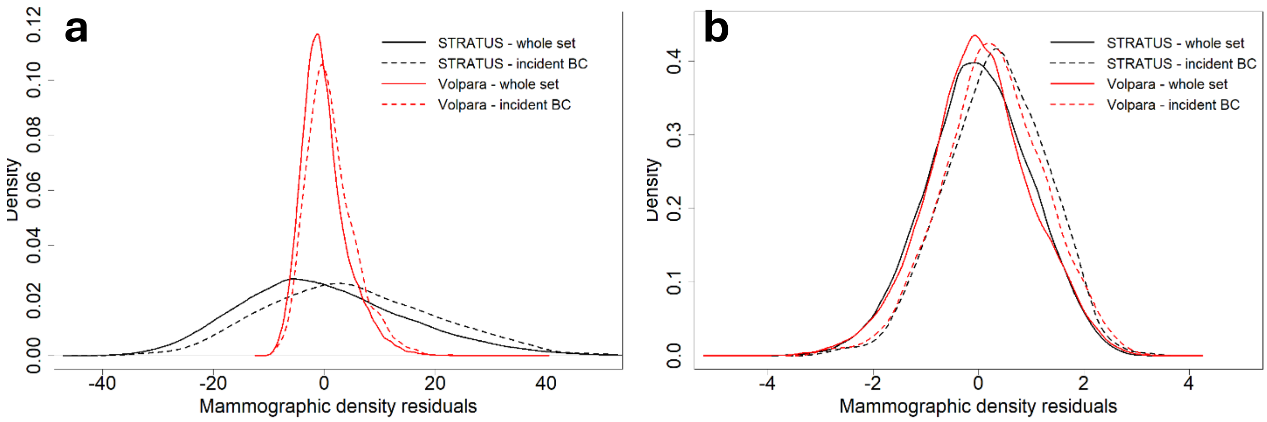

**Figure S1:** Distribution of mammographic density (PMD for STRATUS, PVD for Volpara) residuals after regressing on age and BMI. Panel **a**: residuals before Box-Cox transformation and standardisation. Panel **b**: transformed residuals.

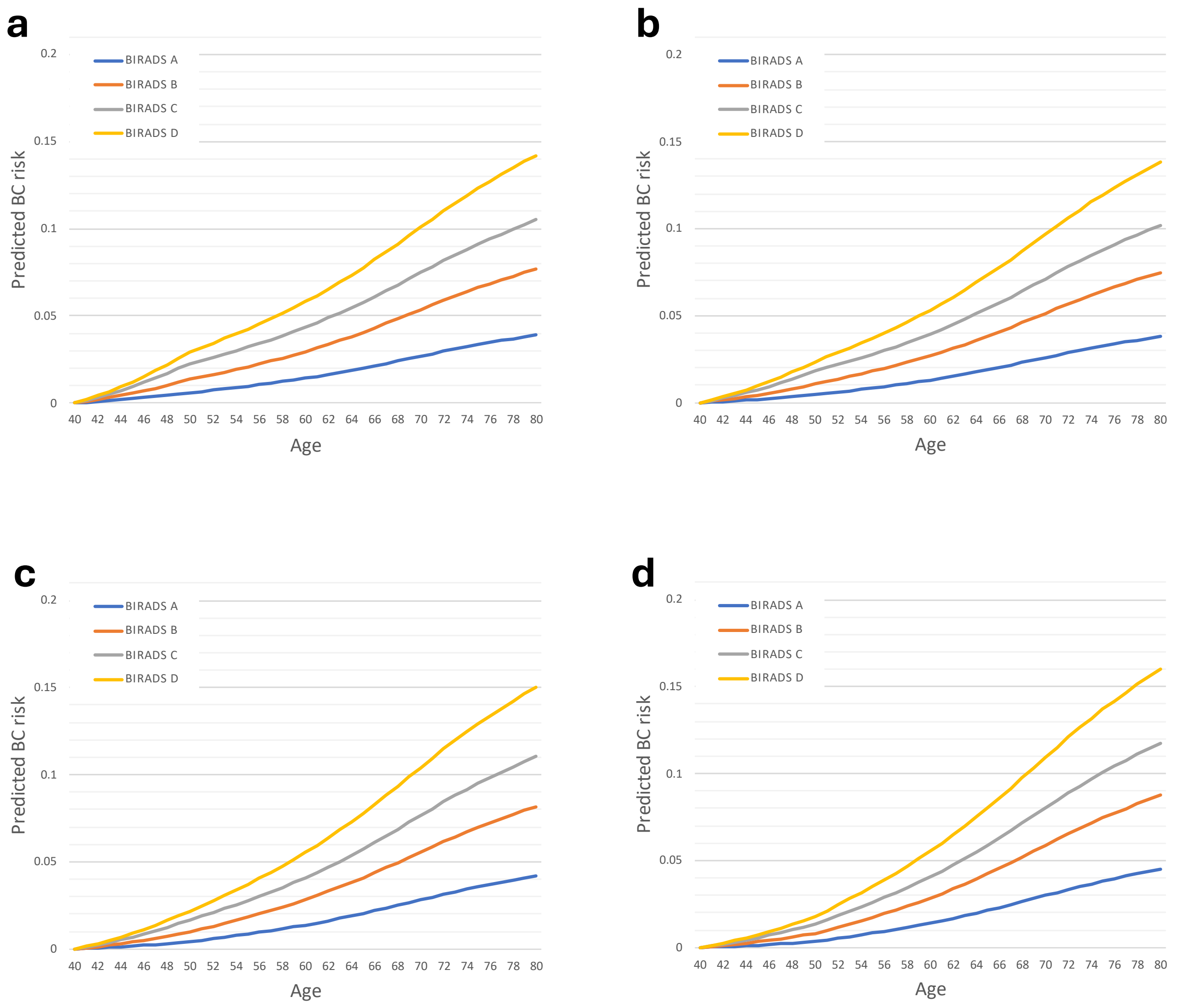

**Figure S2:** Predicted age-specific BC risk for a 40y woman (born in 2000) to age 80y, using the augmented model. MD (as BIRADS categories) and BMI assumed as input information. Panel **a**, BMI < 18.5; panel **b**, BMI in [18.5, 25); panel **c**, BMI in [25, 30); panel **d**, BMI ≥ 30.

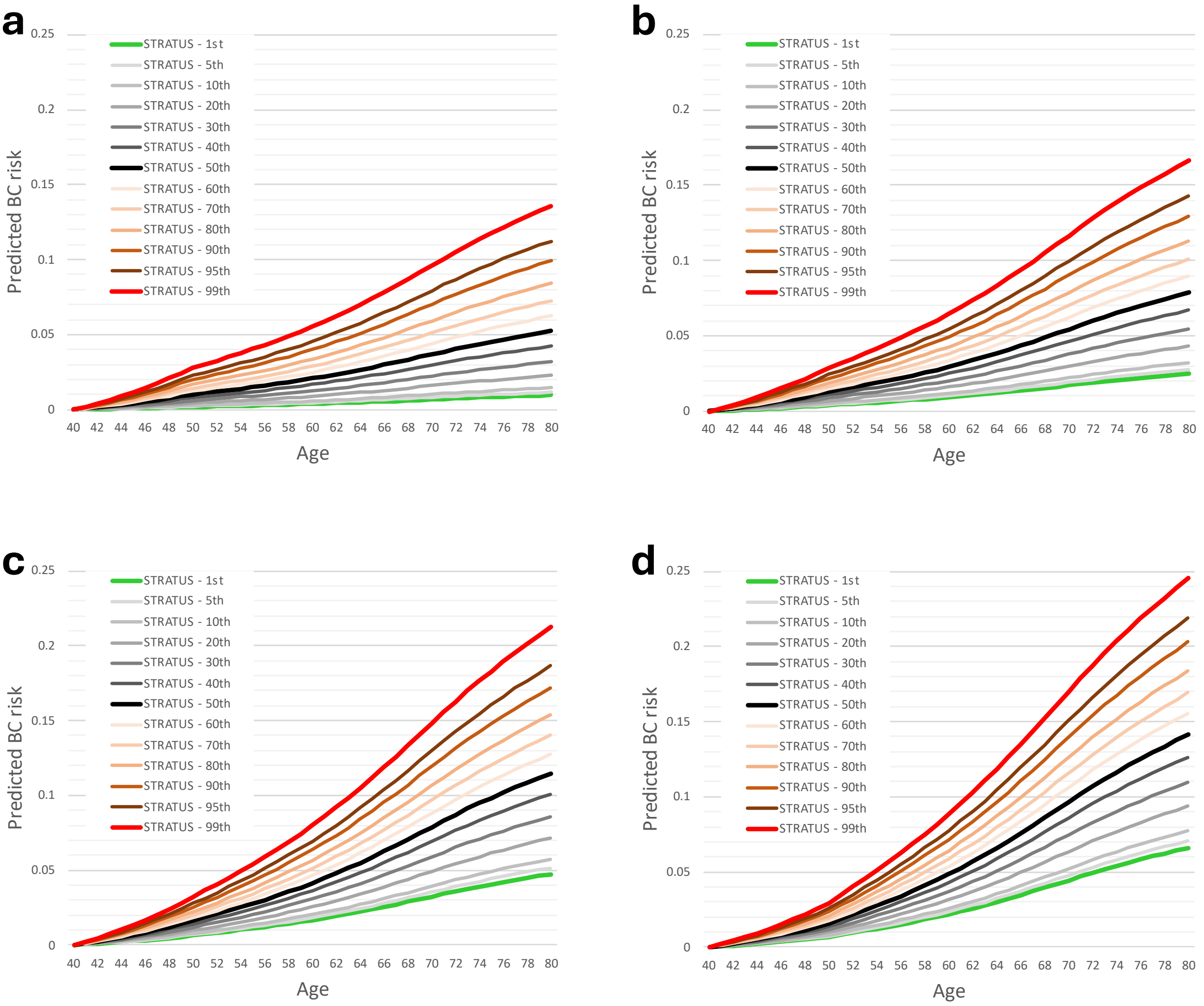

**Figure S3:** Predicted age-specific BC risk for a 40y woman (born in 2000) to age 80y, using the augmented model. MD (as STRATUS PMD) and BMI assumed as input information. Panel **a**, BMI < 18.5; panel **b**, BMI in [18.5, 25); panel **c**, BMI in [25, 30); panel **d**, BMI ≥ 30. Percentiles considered: $1^{st}$,$5^{th}$,$10^{th}$ – $90^{th}$, $95^{th}$, $99^{th}$ of the STRATUS PMD distribution on women <50 years in KARMA.

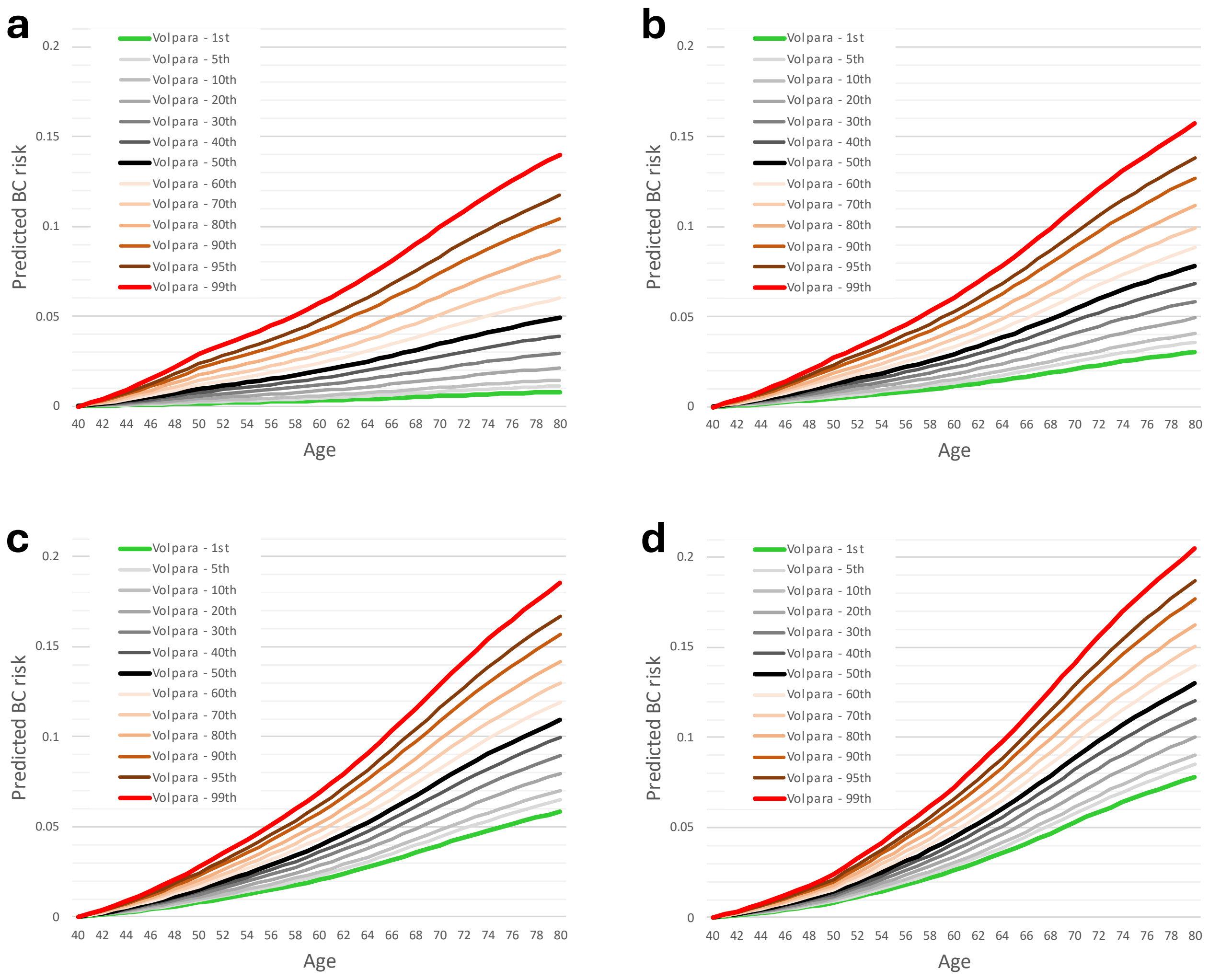

**Figure S4:** Predicted age-specific BC risk for a 40y woman (born in 2000) to age 80y, using the augmented model. MD (as Volpara PVD) and BMI assumed as input information. Panel **a**, BMI < 18.5; panel **b**, BMI in [18.5, 25); panel **c**, BMI in [25, 30); panel **d**, BMI ≥ 30. Percentiles considered: $1^{st}$,$5^{th}$,$10^{th}$ – $90^{th}$, $95^{th}$, $99^{th}$ of the Volpara PVD distribution on women <50 years in KARMA.

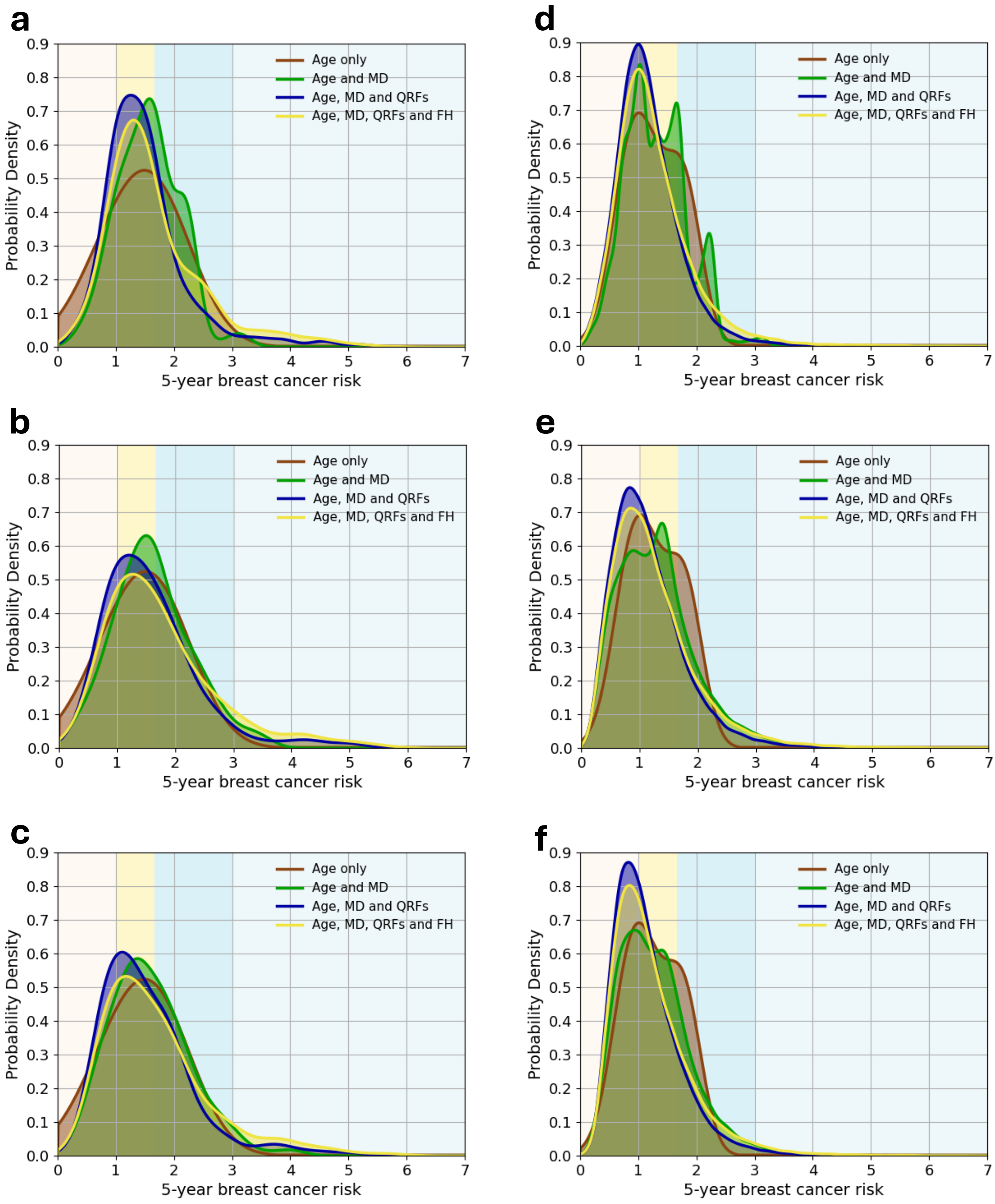

**Figure S5:** Predicted 5-year BC risk based on different combinations of risk predictors: age, mammographic density (MD), questionnaire-based risk factors (QRFs), and family history (FH). (1) women in the testing dataset who developed invasive BC after entering the study (‘affected’, panels **a**, **b**, **c**); (2) unaffected women in the testing dataset (panels **d**, **e**, **f**). All figures show the probability density against the absolute risk: panels (**a**, **d**) using BIRADS categories; panels (**b**, **e**) using STRATUS PMD measures, panels (**c**, **f**) using Volpara PVD measures. The backgrounds of the graphs are shaded to indicate four 5-year BC risk categories: less than 1% (light yellow); between 1% and 1.67% (yellow); between 1.67% and 3% (blue); above 3% (light blue).

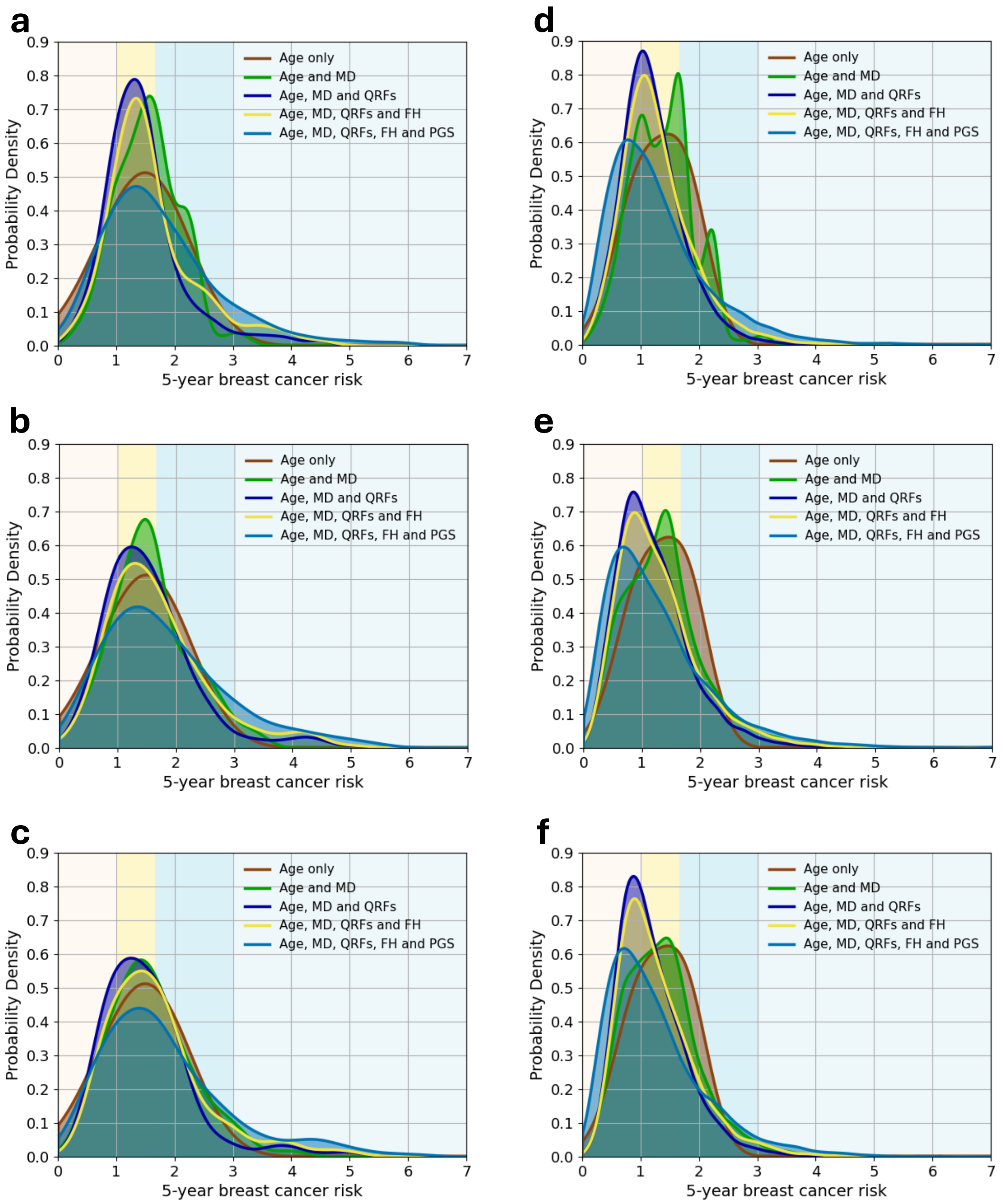

**Figure S6:** Predicted 5-year BC risk based on different combinations of risk predictors: age, mammographic density (MD), questionnaire-based risk factors (QRFs), family history (FH) and polygenic score (PGS). (1) women in the testing subset with PGS data who developed invasive BC after entering the study ('affected', panels **a**, **b**, **c**); (2) unaffected women in the testing subset with PGS data (panels **d**, **e**, **f**). All figures show the probability density against the absolute risk: panels (**a**, **d**) using BIRADS categories, panels (**b**, **e**) using STRATUS PMD measures, panels (**c**, **f**) using Volpara PVD measures. The backgrounds of the graphs are shaded to indicate four risk categories: less than 1% (light yellow); between 1% and 1.67% (yellow); between 1.67% and 3% (blue); above 3% (light blue).

### Tables

**Table S1**: Regression and standardisation parameters used for calculating residuals of PMD (STRATUS) and PVD (Volpara) and then normalising them to achieve a standard normal distribution. Regression required $\lambda_0$, $\lambda_1$, and $\lambda_2$; normalisation required $\lambda_0$, $\lambda_3$, $\mu(X')$, and $\sigma(X')$.

| | **Regression on age at entry and BMI** | | **Regression on age at entry only** | |
|---|---|---|---|---|
| | *STRATUS* | *Volpara* | *STRATUS* | *Volpara* |
| $\lambda_0$ | 89.226 | 25.197 | 69.211 | 19.226 |
| $\lambda_1$ | -0.755 | -0.172 | -0.813 | -0.188 |
| $\lambda_2, BMI < 18.5$ | 0 | 0 | -- | -- |
| $\lambda_2, BMI \in [18.5,25)$ | -15.999 | -4.898 | -- | -- |
| $\lambda_2, BMI \in [25,30)$ | -29.743 | -8.640 | -- | -- |
| $\lambda_2, BMI \geq 30$ | -38.007 | -10.814 | -- | -- |
| $\lambda_3$ | -0.191 | -1.046 | 0.115 | -0.985 |
| $\mu(X')$ | 3.015 | 0.924 | 5.434 | 0.960 |
| $\sigma(X')$ | 0.072 | 0.005 | 0.407 | 0.012 |

**Table S2:** KARMA dataset employed in this study before imputing missing values. Risk factor categories as defined in the BOADICEA model. IQR = interquartile range; SD = standard deviation; FDR = first-degree relatives; BC = breast cancer; HRT = hormone replacement therapy.

| | Full cohort | | Subcohort with PGS | |
|---|---|---|---|---|
| | *Healthy women* | *Incident BC patients* | *Healthy women* | *Incident BC patients* |
| **Number of participants, N** | 60276 | 1167 | 14197 | 682 |
| **Follow-up years, median (IQR)** | 7.6 (7.1-8.1) | 4.2 (2.4-6.1) | 7.5 (7.1-7.9) | 3.2 (2.2-4.3) |
| **Age at baseline, median (IQR)** | 53 (45-62) | 58 (49-66) | 57 (47-65) | 59 (50-66) |
| **Menopausal status, N (%)** | | | | |
| *Pre-menopausal* | 26302 (43.6) | 376 (32.2) | 4969 (35.0) | 205 (30.1) |
| *Post-menopausal* | 14922 (24.8) | 380 (32.6) | 4006 (28.2) | 244 (35.8) |
| *Unknown* | 19052 (31.6) | 411 (35.2) | 5222 (36.8) | 233 (34.2) |
| **Body Mass Index, median (IQR)** | 24.5 (22.3-27.4) | 24.7 (22.5-27.4) | 24.7 (22.4-27.6) | 24.9 (22.6-27.7) |
| **Body Mass Index, N (%)** | | | | |
| *<18.5* | 544 (0.9) | 6 (0.5) | 140 (1.0) | 3 (0.4) |
| *[18.5,25)* | 30946 (51.3) | 591 (50.6) | 7484 (52.7) | 336 (49.3) |
| *[25,30)* | 18020 (29.9) | 373 (32.0) | 4614 (32.5) | 228 (33.4) |
| *≥30* | 7384 (12.3) | 132 (11.3) | 1921 (13.5) | 85 (12.5) |
| *Unknown* | 3382 (5.6) | 65 (5.6) | 38 (0.3) | 30 (4.4) |
| **BIRADS, N (%)** | | | | |
| *A* | 4180 (6.9) | 45 (3.9) | 1099 (7.7) | 28 (4.1) |
| *B* | 21308 (35.4) | 384 (32.9) | 5517 (38.9) | 232 (34.0) |
| *C* | 26254 (43.6) | 540 (46.3) | 5930 (41.8) | 319 (46.8) |
| *D* | 8534 (14.2) | 198 (17.0) | 1651 (11.6) | 103 (15.1) |
| **STRATUS, mean (SD)** | 25.33 (19.36) | 28.04 (19.43) | 23.32 (18.77) | 27.02 (19.08) |
| **Volpara, mean (SD)** | 9.06 (5.22) | 9.64 (5.33) | 8.64 (5.06) | 9.49 (5.23) |
| **Standardised PGS, mean (SD)** | | | 0.006 (1.027) | 0.414 (1.005) |
| **Age at menarche, N (%)** | | | | |
| *<11* | 6284 (10.4) | 123 (10.5) | 725 (5.1) | 66 (9.7) |
| *[11,12)* | 5705 (9.5) | 114 (9.8) | 1425 (10.0) | 58 (8.5) |
| *[12,13)* | 12088 (20.1) | 219 (18.8) | 2966 (20.9) | 128 (18.8) |
| *[13,14)* | 15385 (25.5) | 309 (26.5) | 3775 (26.6) | 186 (27.3) |
| *[14,15)* | 12081 (20.0) | 234 (20.1) | 3065 (21.6) | 147 (21.6) |
| *[15,16)* | 6043 (10.0) | 122 (10.5) | 1571 (11.1) | 69 (10.1) |
| *≥16* | 2690 (4.5) | 46 (3.9) | 670 (4.7) | 28 (4.1) |
| **Parity, N (%)** | | | | |
| *Nulliparous* | 7146 (11.9) | 131 (11.2) | 1739 (12.2) | 73 (10.7) |
| *1 birth* | 8158 (13.5) | 170 (14.6) | 2052 (14.5) | 98 (14.4) |
| *2 births* | 27070 (44.9) | 534 (45.8) | 6731 (47.4) | 332 (48.7) |
| *>2 births* | 13972 (23.2) | 252 (21.6) | 3542 (24.9) | 142 (20.8) |
| *Unknown* | 3930 (6.5) | 80 (6.9) | 133 (0.9) | 37 (5.4) |
| **Age at first live birth, N (%)** | | | | |
| *(no children)* | 7146 (11.9) | 131 (11.2) | 1739 (12.2) | 73 (10.7) |
| *<20* | 2774 (4.6) | 49 (4.2) | 784 (5.5) | 35 (5.1) |
| *[20,25)* | 13578 (22.5) | 266 (22.8) | 3586 (25.3) | 160 (23.5) |
| *[25,30)* | 17403 (28.9) | 345 (29.6) | 4402 (31.0) | 212 (31.1) |
| *≥30* | 15425 (25.6) | 296 (25.4) | 3544 (25.0) | 165 (24.2) |
| *Unknown* | 3950 (6.6) | 80 (6.9) | 142 (1.0) | 37 (5.4) |
| **Alcohol intake g/day, N (%)** | | | | |

| | | | | |
|---|---|---|---|---|
| *0* | 10714 (17.8) | 191 (16.4) | 2669 (18.8) | 112 (16.4) |
| *(0,5)* | 14707 (24.4) | 236 (20.2) | 3641 (25.6) | 145 (21.3) |
| *[5,15)* | 22621 (37.5) | 463 (39.7) | 5681 (39.6) | 279 (40.9) |
| *[15,25)* | 5454 (9.0) | 117 (10.0) | 1440 (10.1) | 76 (11.1) |
| *[25,35)* | 1103 (1.8) | 29 (2.5) | 269 (1.9) | 11 (1.6) |
| *[35,45)* | 1086 (1.8) | 34 (2.9) | 257 (1.8) | 14 (2.1) |
| *≥45* | 205 (0.3) | 7 (0.6) | 56 (0.4) | 4 (0.6) |
| *Unknown* | 4386 (7.3) | 90 (7.7) | 247 (1.7) | 41 (6.0) |
| **Age at menopause, N (%)** | | | | |
| *(no menopause)* | 26302 (43.6) | 376 (32.2) | 4969 (35.0) | 205 (30.1) |
| *<40* | 595 (1.0) | 10 (0.9) | 144 (1.0) | 7 (1.0) |
| *[40,45)* | 1286 (2.1) | 25 (2.1) | 346 (2.4) | 22 (3.2) |
| *[45,50)* | 3681 (6.1) | 85 (7.3) | 944 (7.0) | 61 (8.9) |
| *[50,55)* | 6962 (11.6) | 183 (15.7) | 1815 (12.8) | 107 (15.7) |
| *≥55* | 2398 (4.0) | 77 (6.6) | 707 (5.0) | 47 (6.9) |
| *Unknown* | 19052 (31.6) | 411 (35.2) | 5222 (36.8) | 233 (34.2) |
| **FDR diagnosed with BC (%)** | | | | |
| *0* | 53404 (88.6) | 946 (81.1) | 12468 (87.8) | 559 (82) |
| *1* | 6583 (10.9) | 202 (17.3) | 1660 (11.7) | 116 (17) |
| *≥2* | 289 (0.5) | 19 (1.6) | 69 (0.5) | 7 (1) |
| **Use of oral contraceptives, N (%)** | | | | |
| *Ever* | 47707 (79.1) | 906 (77.6) | 11675 (82.2) | 539 (79.0) |
| *Never* | 8126 (13.5) | 176 (15.1) | 2222 (15.7) | 105 (15.4) |
| *Unknown* | 4443 (7.4) | 85 (7.3) | 300 (2.1) | 38 (5.6) |
| **Use of HRT, N (%)** | | | | |
| *Current, C* | 748 (1.2) | 29 (2.5) | 192 (1.4) | 17 (2.5) |
| *Current, E* | 809 (1.3) | 32 (2.7) | 220 (1.5) | 21 (3.1) |
| *Former* | 7621 (12.6) | 181 (15.5) | 2153 (15.2) | 118 (17.3) |
| *Never* | 43278 (71.8) | 757 (64.9) | 10357 (73.0) | 440 (64.5) |
| *Unknown* | 7820 (13) | 168 (14.4) | 1275 (9.0) | 86 (12.6) |
| **Height (cm), median (IQR)** | 167 (163-170) | 167 (163-171) | 167 (162-170) | 167 (163-171) |

**Table S3:** KARMA dataset employed in this study after imputing missing values. Risk factor categories as defined in the BOADICEA model. IQR = interquartile range; SD = standard deviation; FDR = first-degree relatives; BC = breast cancer; HRT = hormone replacement therapy.

| | **Full cohort** | | **Subcohort with PGS** | |
|---|---|---|---|---|
| | *Healthy women* | *Incident BC patients* | *Healthy women* | *Incident BC patients* |
| **Number of participants, N** | 60276 | 1167 | 14197 | 682 |
| **Follow-up years, median (IQR)** | 7.6 (7.1-8.1) | 4.2 (2.3-6.1) | 7.5 (7.1-7.9) | 3.2 (2.2-4.3) |
| **Age at baseline, median (IQR)** | 53 (45-62) | 58 (49-66) | 57 (47-65) | 59 (50-66) |
| **Menopausal status, N (%)** | | | | |
| *Pre-menopausal* | 32229 (53.5) | 477 (40.9) | 6351 (44.7) | 261 (38.2) |
| *Post-menopausal* | 28047 (46.5) | 690 (59.1) | 7846 (55.3) | 421 (61.8) |
| **Body Mass Index ($kg/m^2$), median (IQR)** | 24.6 (22.3-27.5) | 24.7 (22.5-27.5) | 24.7 (22.4-27.5) | 24.9 (22.6-27.7) |
| **Body Mass Index, N (%)** | | | | |
| *<18.5* | 568 (0.9) | 7 (0.6) | 141 (1.0) | 3 (0.4) |
| *[18.5,25)* | 32486 (53.9) | 620 (53.1) | 7505 (52.9) | 349 (51.2) |
| *[25,30)* | 19157 (31.8) | 396 (33.9) | 4623 (32.5) | 238 (34.9) |
| *≥30* | 8066 (13.4) | 144 (12.3) | 1928 (13.6) | 92 (13.5) |
| **BIRADS, N (%)** | | | | |
| *A* | 4180 (6.9) | 45 (3.9) | 1099 (7.7) | 28 (4.1) |
| *B* | 21308 (35.4) | 384 (32.9) | 5517 (38.9) | 232 (34.0) |
| *C* | 26254 (43.6) | 540 (46.3) | 5930 (41.8) | 319 (46.8) |
| *D* | 8534 (14.2) | 198 (17.0) | 1651 (11.6) | 103 (15.1) |
| **STRATUS res., mean (SD)** | 0.04 (15.28) | 5.16 (15.43) | -0.10 (14.92) | 5.08 (14.78) |
| **Volpara res., mean (SD)** | 0.00 (4.25) | 1.15 (4.42) | 0.02 (4.13) | 1.23 (4.25) |
| **Standardised PGS, mean (SD)** | | | 0.006 (1.027) | 0.414 (1.005) |
| **Age at menarche, N (%)** | | | | |
| *<11* | 6284 (10.4) | 123 (10.5) | 725 (5.1) | 66 (9.7) |
| *[11,12)* | 5705 (9.5) | 114 (9.8) | 1425 (10.0) | 58 (8.5) |
| *[12,13)* | 12088 (20.1) | 219 (18.8) | 2966 (20.9) | 128 (18.8) |
| *[13,14)* | 15385 (25.5) | 309 (26.5) | 3775 (26.6) | 186 (27.3) |
| *[14,15)* | 12081 (20.0) | 234 (20.1) | 3065 (21.6) | 147 (21.6) |
| *[15,16)* | 6043 (10.0) | 122 (10.5) | 1571 (11.1) | 69 (10.1) |
| *≥16* | 2690 (4.5) | 46 (3.9) | 670 (4.7) | 28 (4.1) |
| **Parity, N (%)** | | | | |
| *Nulliparous* | 7686 (12.8) | 142 (12.8) | 1756 (12.3) | 78 (11.4) |
| *1 birth* | 8754 (14.5) | 184 (15.8) | 2073 (14.6) | 106 (15.5) |
| *2 births* | 28946 (48.0) | 572 (49.1) | 6793 (47.9) | 349 (51.2) |
| *>2 births* | 14889 (24.7) | 268 (23.0) | 3572 (25.2) | 149 (21.8) |
| **Age at first live birth, N (%)** | | | | |
| *(no children)* | 7688 (12.8) | 142 (12.2) | 1756 (12.4) | 78 (11.4) |
| *<20* | 2958 (4.9) | 53 (4.5) | 793 (5.6) | 38 (5.6) |
| *[20,25)* | 14458 (24.0) | 286 (24.5) | 3624 (25.6) | 170 (24.9) |
| *[25,30)* | 18596 (30.9) | 368 (31.5) | 4444 (31.3) | 222 (32.6) |
| *≥30* | 16577 (27.4) | 318 (27.2) | 3579 (25.2) | 174 (25.5) |
| **Alcohol intake g/day, N (%)** | | | | |
| *0* | 11619 (19.3) | 206 (17.7) | 2720 (19.2) | 120 (17.6) |
| *(0,5)* | 15862 (26.3) | 260 (22.3) | 3706 (26.1) | 156 (22.9) |
| *[5,15)* | 24348 (40.4) | 500 (42.8) | 5716 (40.3) | 295 (43.3) |
| *[15,25)* | 5868 (9.7) | 127 (10.9) | 1463 (10.3) | 80 (11.7) |

| | | | | |
|---|---|---|---|---|
| *[25,35)* | 1186 (2.0) | 31 (2.7) | 273 (1.9) | 12 (1.8) |
| *[35,45)* | 1171 (1.9) | 36 (3.1) | 261 (1.8) | 15 (2.2) |
| *≥45* | 222 (0.4) | 7 (0.6) | 57 (0.4) | 4 (0.6) |
| **Age at menopause, N (%)** | | | | |
| *(no menopause)* | 32229 (53.5) | 477 (40.9) | 6351 (44.7) | 261 (38.2) |
| *<40* | 1142 (1.9) | 23 (2.0) | 286 (2.0) | 14 (2.0) |
| *[40,45)* | 2408 (4.0) | 48 (4.1) | 631 (4.4) | 36 (5.3) |
| *[45,50)* | 6548 (10.9) | 150 (12.9) | 1779 (12.2) | 98 (14.3) |
| *[50,55)* | 12950 (21.5) | 329 (28.2) | 3632 (25.6) | 190 (28.0) |
| *≥55* | 4999(8.3) | 140 (12.0) | 1518 (10.7) | 83 (12.2) |
| **FDR diagnosed with BC (%)** | | | | |
| *0* | 53404 (88.6) | 946 (81.1) | 12468 (87.8) | 559 (82) |
| *1* | 6583 (10.9) | 202 (17.3) | 1660 (11.7) | 116 (17) |
| *≥2* | 289 (0.5) | 19 (1.6) | 69 (0.5) | 7 (1) |
| **Use of oral contraceptives, N (%)** | | | | |
| *Ever* | 51544 (85.5) | 976 (83.6) | 11914 (83.9) | 569 (83.4) |
| *Never* | 8732 (14.5) | 191 (16.4) | 2283 (16.1) | 113 (16.6) |
| **Use of HRT, N (%)** | | | | |
| *Current, C* | 878 (1.5) | 36 (3.1) | 218 (1.5) | 21 (3.1) |
| *Current, E* | 949 (1.6) | 36 (3.1) | 250 (1.8) | 23 (3.4) |
| *Former* | 8993 (14.9) | 213 (18.2) | 2440 (17.2) | 136 (19.9) |
| *Never* | 49456 (82.0) | 883 (75.6) | 11290 (79.5) | 502 (73.6) |
| **Height (cm), median (IQR)** | 167 (162-170) | 167 (163-171) | 167 (162-170) | 167 (163-171) |

**Table S4:** Training dataset, randomly extracted (*R* seed= 24) from the imputed KARMA dataset (table S3). Risk factor categories as defined in the BOADICEA model. IQR = interquartile range; SD = standard deviation; FDR = first-degree relatives; BC = breast cancer; HRT = hormone replacement therapy.

| | **Full cohort** | | **Subcohort with PGS** | |
|---|---|---|---|---|
| | *Healthy women* | *Incident BC patients* | *Healthy women* | *Incident BC patients* |
| **Number of participants, N** | 40143 | 777 | 9445 | 468 |
| **Follow-up years, median (IQR)** | 7.6 (7.1-8.1) | 4.2 (2.3-6.1) | 7.5 (7.1-7.9) | 3.2 (2.2-4.2) |
| **Age at baseline, median (IQR)** | 53 (45-62) | 58 (48-66) | 57 (48-65) | 59 (50-66) |
| **Menopausal status, N (%)** | | | | |
| *Pre-menopausal* | 21419 (53.4) | 335 (43.1) | 4185 (44.3) | 185 (39.5) |
| *Post-menopausal* | 18724 (46.6) | 442 (56.9) | 5260 (55.7) | 283 (60.5) |
| **Body Mass Index (kg/m$^2$), median (IQR)** | 24.6 (22.3-27.5) | 24.8 (22.5-27.5) | 24.7 (22.4-27.6) | 24.8 (22.5-27.7) |
| **Body Mass Index, N (%)** | | | | |
| *<18.5* | 370 (0.9) | 5 (0.6) | 92 (1.0) | 2 (0.4) |
| *[18.5,25)* | 21538 (53.7) | 405 (52.2) | 4966 (52.6) | 243 (51.9) |
| *[25,30)* | 12836 (32.0) | 280 (36.1) | 3096 (32.8) | 169 (36.1) |
| *≥30* | 5399 (13.4) | 86 (11.1) | 1290 (13.7) | 54 (11.5) |
| **BIRADS, N (%)** | | | | |
| *A* | 2779 (6.9) | 25 (3.2) | 750 (7.9) | 16 (3.4) |
| *B* | 14253 (35.5) | 261 (33.6) | 3684 (39.0) | 160 (34.2) |
| *C* | 17429 (43.4) | 356 (45.8) | 3908 (41.4) | 216 (46.2) |
| *D* | 5682 (14.2) | 135 (17.4) | 1103 (11.7) | 76 (16.2) |
| **STRATUS res., mean (SD)** | 0.04 (15.25) | 5.27 (15.63) | -0.09 (14.92) | 5.47 (15.01) |
| **Volpara res., mean (SD)** | 0.01 (4.24) | 1.12 (4.43) | 0.05 (4.13) | 1.26 (4.25) |
| **Standardised PGS, mean (SD)** | | | 0.011 (1.037) | 0.445 (1.041) |
| **Age at menarche, N (%)** | | | | |
| *<11* | 4173 (10.4) | 79 (10.2) | 478 (5.1) | 45 (9.6) |
| *[11,12)* | 3758 (9.4) | 70 (9.0) | 940 (10.0) | 41 (8.8) |
| *[12,13)* | 8111 (20.2) | 147 (18.9) | 1990 (21.1) | 89 (19.0) |
| *[13,14)* | 10211 (25.4) | 207 (26.6) | 2491 (26.4) | 120 (25.6) |
| *[14,15)* | 8073 (20.1) | 160 (20.6) | 2074 (22.0) | 102 (21.8) |
| *[15,16)* | 4072 (10.1) | 82 (10.6) | 1047 (11.1) | 49 (10.5) |
| *≥16* | 1745 (4.3) | 32 (4.1) | 425 (4.5) | 22 (4.7) |
| **Parity, N (%)** | | | | |
| *Nulliparous* | 5155 (12.8) | 94 (12.1) | 1189 (12.6) | 50 (10.7) |
| *1 birth* | 5786 (14.4) | 118 (15.2) | 1351 (14.3) | 66 (14.1) |
| *2 births* | 19314 (48.1) | 382 (49.2) | 4537 (48.0) | 249 (53.3) |
| *>2 births* | 9888 (24.6) | 182 (23.5) | 2368 (25.1) | 102 (21.8) |
| **Age at first live birth, N (%)** | | | | |
| *(no children)* | 5157 (12.8) | 94 (12.1) | 1189 (12.6) | 50 (10.7) |
| *<20* | 1979 (4.9) | 39 (5.0) | 518 (5.5) | 28 (6.0) |
| *[20,25)* | 9624 (24.0) | 196 (25.3) | 2409 (25.5) | 124 (26.5) |
| *[25,30)* | 12342 (30.7) | 250 (32.2) | 2954 (31.3) | 155 (33.1) |
| *≥30* | 11041 (27.5) | 197 (25.4) | 2375 (25.1) | 111 (23.7) |
| **Alcohol intake g/day, N (%)** | | | | |
| *0* | 7738 (19.3) | 139 (17.9) | 1788 (18.9) | 80 (17.1) |
| *(0,5)* | 10579 (26.4) | 173 (22.3) | 2460 (26.0) | 113 (24.1) |
| *[5,15)* | 16187 (40.3) | 331 (42.6) | 3838 (40.6) | 199 (42.5) |
| *[15,25)* | 3898 (9.7) | 81 (10.4) | 968 (10.2) | 54 (11.5) |

| | | | | |
|---|---|---|---|---|
| *[25,35)* | 781 (1.9) | 22 (2.8) | 175 (1.9) | 8 (1.7) |
| *[35,45)* | 814 (2.0) | 27 (3.5) | 180 (1.9) | 11 (2.4) |
| *≥45* | 146 (0.4) | 4 (0.5) | 36 (0.4) | 3 (0.6) |
| **Age at menopause, N (%)** | | | | |
| *(no menopause)* | 21419 (53.4) | 335 (43.1) | 4185 (44.3) | 185 (39.5) |
| *<40* | 758 (1.9) | 14 (1.8) | 184 (1.9) | 9 (1.9) |
| *[40,45)* | 1606 (4.0) | 31 (4.0) | 426 (4.5) | 23 (4.9) |
| *[45,50)* | 4347 (10.8) | 87 (12.5) | 1186 (12.6) | 66 (14.1) |
| *[50,55)* | 8688 (21.6) | 212 (27.3) | 2456 (26.0) | 127 (27.1) |
| *≥55* | 3325 (8.3) | 88 (11.3) | 1008 (10.7) | 58 (12.4) |
| **FDR diagnosed with BC (%)** | | | | |
| *0* | 35610 (88.7) | 633 (81.5) | 8306 (87.9) | 386 (82.5) |
| *1* | 4358 (10.9) | 135 (17.4) | 1088 (11.5) | 79 (16.9) |
| *≥2* | 175 (0.4) | 9 (1.2) | 51 (0.5) | 3 (0.6) |
| **Use of oral contraceptives, N (%)** | | | | |
| *Ever* | 34331 (85.5) | 646 (83.0) | 7953 (84.2) | 388 (82.9) |
| *Never* | 5812 (14.5) | 132 (17.0) | 1492 (15.8) | 80 (17.1) |
| **Use of HRT, N (%)** | | | | |
| *Current, C* | 598 (1.5) | 26 (3.3) | 160 (1.7) | 17 (3.6) |
| *Current, E* | 618 (1.5) | 18 (2.3) | 162 (1.7) | 11 (2.4) |
| *Former* | 6014 (15.0) | 126 (16.2) | 1619 (17.1) | 84 (17.9) |
| *Never* | 32913 (82.0) | 608 (78.1) | 7504 (79.4) | 356 (76.1) |
| **Height (cm), median (IQR)** | 167 (162-170) | 167 (163-171) | 167 (162-170) | 167 (163-171) |

**Table S5:** Testing dataset, randomly extracted (*R* seed= 24) from the imputed KARMA dataset (table S3). Risk factor categories as defined in the BOADICEA model. IQR = interquartile range; SD = standard deviation; FDR = first-degree relatives; BC = breast cancer; HRT = hormone replacement therapy.

| | **Full cohort** | | **Subcohort with PGS** | |
|---|---|---|---|---|
| | *Healthy women* | *Incident BC patients* | *Healthy women* | *Incident BC patients* |
| **Number of participants, N** | 20133 | 390 | 4752 | 214 |
| **Follow-up years, median (IQR)** | 7.6 (7.1-8.1) | 4.4 (2.4-6.2) | 7.5 (7.1-7.9) | 3.2 (2.2-4.4) |
| **Age at baseline, median (IQR)** | 53 (45-62) | 59 (50-66) | 56 (47-65) | 59 (50-66) |
| **Menopausal status, N (%)** | | | | |
| *Pre-menopausal* | 10809 (53.7) | 142 (36.4) | 2166 (45.6) | 76 (35.5) |
| *Post-menopausal* | 9324 (46.3) | 248 (63.6) | 2586 (54.4) | 138 (64.5) |
| **Body Mass Index (kg/m$^2$), median (IQR)** | 24.5 (22.3-27.4) | 24.7 (22.6-27.4) | 24.6 (22.3-27.5) | 25.0 (23.0-27.8) |
| **Body Mass Index, N (%)** | | | | |
| *<18.5* | 198 (1.0) | 1 (0.3) | 48 (1.0) | 1 (0.5) |
| *[18.5,25)* | 10948 (54.4) | 215 (55.1) | 2539 (53.4) | 106 (49.5) |
| *[25,30)* | 6321 (31.4) | 116 (29.7) | 1526 (32.1) | 69 (32.2) |
| *≥30* | 2667 (13.2) | 58 (14.9) | 639 (13.4) | 38 (17.8) |
| **BIRADS, N (%)** | | | | |
| *A* | 1401 (7.0) | 20 (5.1) | 349 (7.3) | 12 (5.6) |
| *B* | 7055 (35.0) | 123 (31.5) | 1833 (38.6) | 72 (33.6) |
| *C* | 8825 (43.8) | 184 (47.2) | 2022 (42.6) | 103 (48.1) |
| *D* | 2852 (14.2) | 63 (16.2) | 548 (11.5) | 27 (12.6) |
| **STRATUS res., mean (SD)** | 0.03 (15.33) | 4.94 (15.04) | -0.11 (14.92) | 4.21 (14.24) |
| **Volpara res., mean (SD)** | -0.03 (4.25) | 1.20 (4.41) | -0.04 (4.14) | 1.15 (4.26) |
| **Standardised PGS, mean (SD)** | | | -0.005 (1.007) | 0.345 (0.919) |
| **Age at menarche, N (%)** | | | | |
| *<11* | 2111 (10.5) | 44 (11.3) | 247 (5.2) | 21 (9.8) |
| *[11,12)* | 1947 (9.7) | 44 (11.3) | 485 (10.2) | 17 (7.9) |
| *[12,13)* | 3977 (19.8) | 72 (18.5) | 976 (20.5) | 39 (18.2) |
| *[13,14)* | 5174 (25.7) | 102 (26.5) | 1284 (27.0) | 66 (30.9) |
| *[14,15)* | 4008 (19.9) | 74 (19.0) | 991 (20.9) | 45 (21.0) |
| *[15,16)* | 1972 (9.8) | 40 (10.3) | 524 (11.0) | 20 (9.3) |
| *≥16* | 945 (4.7) | 14 (3.6) | 245 (5.2) | 6 (2.8) |
| **Parity, N (%)** | | | | |
| *Nulliparous* | 2531 (12.6) | 48 (12.3) | 568 (12.0) | 28 (13.1) |
| *1 birth* | 2968 (14.7) | 66 (17.0) | 722 (15.2) | 39 (18.2) |
| *2 births* | 9632 (47.8) | 189 (48.6) | 2256 (47.5) | 100 (46.7) |
| *>2 births* | 5002 (24.8) | 86 (22.1) | 1207 (25.4) | 47 (22.0) |
| **Age at first live birth, N (%)** | | | | |
| *(no children)* | 2531 (12.6) | 48 (12.3) | 568 (12.0) | 28 (13.1) |
| *<20* | 978 (4.9) | 14 (3.6) | 275 (5.8) | 10 (4.7) |
| *[20,25)* | 4834 (24.0) | 90 (23.1) | 1215 (25.6) | 46 (21.5) |
| *[25,30)* | 6254 (31.1) | 117 (30.0) | 1490 (31.4) | 67 (31.3) |
| *≥30* | 5536 (27.5) | 121 (31.0) | 1204 (25.3) | 63 (29.4) |
| **Alcohol intake g/day, N (%)** | | | | |
| *0* | 3881 (19.3) | 67 (17.2) | 932 (19.6) | 40 (18.8) |
| *(0,5)* | 5282 (26.2) | 87 (22.4) | 1246 (26.2) | 43 (20.1) |
| *[5,15)* | 8161 (40.5) | 169 (43.4) | 1878 (39.5) | 96 (45.1) |
| *[15,25)* | 1970 (9.8) | 46 (11.8) | 496 (10.4) | 25 (11.7) |

| | | | | |
|---|---|---|---|---|
| *[25,35)* | 405 (2.0) | 9 (2.3) | 99 (2.1) | 4 (1.9) |
| *[35,45)* | 357 (1.8) | 8 (2.1) | 80 (1.7) | 4 (1.9) |
| *≥45* | 76 (0.4) | 3 (0.8) | 21 (0.4) | 1 (0.5) |
| **Age at menopause, N (%)** | | | | |
| *(no menopause)* | 10809 (53.7) | 142 (36.4) | 2166 (45.6) | 76 (35.5) |
| *<40* | 384 (1.9) | 8 (2.1) | 102 (2.1) | 4 (1.9) |
| *[40,45)* | 803 (4.0) | 18 (4.6) | 205 (4.3) | 13 (6.1) |
| *[45,50)* | 2201 (10.9) | 53 (13.6) | 592 (12.5) | 32 (15.0) |
| *[50,55)* | 4262 (21.2) | 117 (30.0) | 1176 (24.8) | 64 (29.9) |
| *≥55* | 1674(8.3) | 52 (13.3) | 510 (10.7) | 25 (11.7) |
| **FDR diagnosed with BC (%)** | | | | |
| *0* | 17794 (88.4) | 313 (80.3) | 4162 (87.6) | 173 (80.8) |
| *1* | 2225 (11.1) | 67 (17.2) | 572 (12.0) | 37 (17.3) |
| *≥2* | 114 (0.6) | 10 (2.6) | 18 (0.4) | 4 (1.9) |
| **Use of oral contraceptives, N (%)** | | | | |
| *Ever* | 17212 (85.5) | 331 (84.9) | 3961 (83.4) | 181 (84.6) |
| *Never* | 2920 (14.5) | 59 (15.1) | 791 (16.6) | 33 (15.4) |
| **Use of HRT, N (%)** | | | | |
| *Current, C* | 280 (1.4) | 10 (2.6) | 58 (1.2) | 4 (1.9) |
| *Current, E* | 332 (1.6) | 18 (4.6) | 87 (1.8) | 12 (5.6) |
| *Former* | 2978 (14.8) | 87 (22.3) | 821 (17.3) | 52 (24.3) |
| *Never* | 16543 (82.2) | 275 (70.5) | 3786 (79.7) | 146 (68.2) |
| **Height (cm), median (IQR)** | 167 (163-170) | 166 (163-170) | 167 (163-170) | 167 (163-171) |

**Table S6:** Hazard ratio estimates per standard deviation of the standardised STRATUS PMD and Volpara VPD residuals; obtained by regressing on age, and then by adjusting for FH and QRFs (BMI included).

| | **STRATUS** | | | **Volpara** | | |
|---|---|---|---|---|---|---|
| | *Unknown status* | *Pre-menopausal* | *Post-menopausal* | *Unknown status* | *Pre-menopausal* | *Post-menopausal* |
| *Unadjusted for PGS* | 1.54 (1.42-1.68) | 1.58 (1.41-1.77) | 1.51 (1.34-1.70) | 1.44 (1.31-1.57) | 1.38 (1.24-1.54) | 1.51 (1.33-1.73) |
| *Adjusted for PGS* | 1.51 (1.39-1.65) | 1.55 (1.39-1.72) | 1.48 (1.33-1.66) | 1.42 (1.30-1.55) | 1.37 (1.23-1.53) | 1.50 (1.32-1.71) |

**Table S7:** Summary of the reclassification towards lower, identical, or higher categories when comparing risks predicted with the original BOADICEA v7 model vs the augmented model (using BIRADS categories, STRATUS PMD and Volpara PVD as MD inputs. Thresholds of risk categories derived from Tice *et al.*, 2015[13]. Risk distributions obtained with models (1) considering MD only; (2) considering MD, FH, QRFs and PGS. All models include age by default.

| | **All entries** | | | **Unaffected only** | | | **Affected only** | | |
|---|---|---|---|---|---|---|---|---|---|
| **Considering MD only** | *lower* | *same* | *higher* | *lower* | *same* | *higher* | *lower* | *same* | *higher* |
| *new model (BIRADS)* | 0.000 | 0.871 | 0.129 | 0.000 | 0.872 | 0.128 | 0.000 | 0.834 | 0.166 |
| *new model (STRATUS)* | 0.111 | 0.705 | 0.183 | 0.112 | 0.705 | 0.183 | 0.061 | 0.710 | 0.229 |
| *new model (Volpara)* | 0.122 | 0.701 | 0.177 | 0.122 | 0.702 | 0.176 | 0.103 | 0.666 | 0.231 |
| | **All entries** | | | **Unaffected only** | | | **Affected only** | | |
| **Considering full model** | *lower* | *same* | *higher* | *lower* | *same* | *higher* | *lower* | *same* | *higher* |
| *new model (BIRADS)* | 0.000 | 0.899 | 0.101 | 0.000 | 0.899 | 0.101 | 0.000 | 0.878 | 0.122 |
| *new model (STRATUS)* | 0.082 | 0.778 | 0.140 | 0.082 | 0.779 | 0.139 | 0.059 | 0.702 | 0.239 |
| *new model (Volpara)* | 0.097 | 0.767 | 0.136 | 0.097 | 0.768 | 0.135 | 0.107 | 0.660 | 0.232 |

**Table S8:** Hazard ratio estimates for a woman with STRATUS PMD or Volpara VPD 1SD higher than the population average (in the KARMA cohort); calculated for different ages and BMI categories. Women of age 40y and 50y were assumed to be pre-menopausal; women of age 60y and 70y were assumed to be post-menopausal.

| | STRATUS | | | | Volpara | | | |
|---|---|---|---|---|---|---|---|---|
| | *Age 40y* | *Age 50y* | *Age 60y* | *Age 70y* | *Age 40y* | *Age 50y* | *Age 60y* | *Age 70y* |
| *BMI 1°* $< 18.5$ | 0.69 | 0.86 | 1.04 | 1.22 | 0.78 | 0.88 | 0.99 | 1.14 |
| *BMI 2°* $\in [18.5,25)$ | 1.07 | 1.28 | 1.44 | 1.64 | 1.08 | 1.19 | 1.40 | 1.54 |
| *BMI 3°* $\in [25,30)$ | 1.47 | 1.71 | 1.81 | 2.04 | 1.30 | 1.40 | 1.71 | 1.84 |
| *BMI 4°* $\geq 30$ | 1.74 | 2.00 | 2.06 | 2.29 | 1.42 | 1.51 | 1.88 | 2.01 |
| *BMI unknown* | 1.29 | 1.55 | 1.73 | 1.99 | 1.26 | 1.38 | 1.68 | 1.84 |
